# Advancements in Managing Wound Biofilm: A Systematic Review and Meta-Analysis of Randomized Controlled Trials on Topical Modalities

**DOI:** 10.1101/2023.08.20.23294342

**Authors:** Adam Astrada, Rian Adi Pamungkas, Khoirul Rista Abidin M. Biomed

**Author notes:** corresponding author: Adam Astrada, DHSc., CNS, RN Assistant Professor School of Nursing, Faculty of Health Sciences, Esa Unggul University, Indonesia Jl. Arjuna Utara No.9, Kebon Jeruk, Jakarta, Indonesia 11510.

## Abstract

**Significance:** Gaining insights into the efficacy of topical wound treatments for biofilm eradication is pivotal to enhancing wound management strategies. Despite numerous available agents claiming anti-biofilm properties, the substantiating evidence remains inconclusive. This study aimed to assess the immediate impact of topical wound treatments on wound biofilm and healing outcomes in acute and chronic ulcers.

**Recent Advances:** We comprehensively searched PubMed, ClinicalTrials.gov, and Google Scholar. Additionally, eligible grey literature was incorporated. English-language randomized controlled trials, observational, cohort, and case-control studies targeting biofilm prevention, inhibition, or elimination across diverse wound types were included. Primary outcomes included biofilm presence and elimination, supplemented by secondary outcomes encompassing reduced wound size, complete closure, and diminished infection indicators. Bacterial load reduction and biofilm presence were also assessed.

**Critical Issues:** Twenty-eight articles met the inclusion criteria. Various modalities were identified, including biofilm-visualization techniques like wound blotting and handheld autofluorescence imaging. A notable high bias risk was observed across all studies. Pooled analysis for the primary outcomes was infeasible due to limited eligible studies and data reporting challenges. The primary source of bias is the lack of consensus on objectively evaluating the biofilm. As for the secondary outcomes, the pooled analysis for complete wound closure (6 RCTs, yielding n=439) and presence of infection/inflammation (2 RCTs, yielding n=284) showed no significant difference, with log odds ratio (LOD) 0.03 (95% CI: – 1.02 – 1.09; τ^2^=1.09, SE: 1.10, Q=14.33, *p*=.014) and LOD – 0.95 (95% CI: – 3.54 – 1.64; τ^2^=2.32, Q=2.71, *p*=.10), respectively.

**Future Direction:** Our findings suggest insufficient evidence to support anti-biofilm claims of topical modalities. Clinicians’ skill appears to play a pivotal role in biofilm elimination and wound healing enhancement, with potential optimization through visual-guided techniques like wound blotting and autofluorescence imaging. More rigorous clinical trials are warranted to ascertain the efficacy of these techniques.

## 1. Scope and Significance

Various products and approaches in the market claim to have antibiofilm effects, eventually improving wound healing. Yet, the studies regarding this claim are inconclusive. This study aimed to investigate the pooled effect of topical wound treatments on wound biofilm and healing outcomes in acute and chronic ulcers.

## 2. Translational Relevance

This review found insufficient evidence to conclude that any topical modalities, namely dressings, ointments, and wound bed preparation techniques, could reduce wound surface biofilm across studies. The problem may be raised from the need for more consensus on the objective evaluation of biofilm itself.

## 3. Clinical Relevance

The result further emphasizes the critical role of wound care clinicians’ skills in wound biofilm management instead of the types of products or topical modalities used.

## 4. Background

### Rationale

Biofilm has become a growing health concern since it contributes to 80% of human infections.^1^ Microorganisms living within a biofilm are up to 1,000 times more antibiotic-resistant than their planktonic forms.^2^ Biofilm attached to indwelled devices, such as implants, can cause early implant removal and surgical site infections. The absence of vascularization in implants creates a potential dead space that fosters microbial attachment and biofilm development. Biofilm-associated costs in wounds and surgery are estimated to reach several 3.3 billion US dollars in European countries.^3^ Moreover, a growing health threat by superbugs and multi-drug-resistant pathogens amplifies the need for therapeutic modalities to tackle biofilms, especially those associated with wounds.

It is estimated that 78.2% of wounds present biofilms.^4^ Since biofilm is ubiquitous in wounds, multi-modal approaches have been proposed and developed to combat this situation. More than 40 agents available on the market in various forms claim to have anti-biofilm effects.^5^

Yet, the evidence supporting this claim remains unclear. Therefore, this systematic review and meta-analysis aimed to provide pooled data on topical wound treatments such as cleansers, ointment, dressings, and therapeutic modalities and their immediate effects on biofilm elimination and wound healing in acute and chronic ulcers.

## 5. Methods

The search strategy was based on the research question of “To patients with acute or chronic ulcers (participants), do the topical wound treatments such as cleansers, ointment, dressings, and therapeutic modalities (intervention) provide anti-biofilm effects or eliminate biofilms compared to the standard of care or among the products (comparators).” The review protocol is reported according to Preferred Reporting Items for Systematic Reviews and Meta-Analyses (PRISMA) 2020 and registered in PROSPERO 2023 (ID: CRD42023407421), available from: https://www.crd.york.ac.uk/prospero/display_record.php?ID=CRD42023407421

### 5.1 Eligibility Criteria

We aimed to find, assess and synthesize all randomized controlled trials, observational studies (all types), cohort (longitudinal) studies, or case-control studies containing all types of wounds (e.g., acute and chronic wounds, such as diabetic foot ulcers, venous ulcers, surgical site infections, etc.) or clinical studies and trials (e.g., randomized controlled trials, non-controlled interventional studies, observational studies involving human subjects). If they used the following interventions wound dressings, ointments, or techniques purposed to prevent, inhibit, or eliminate wound biofilms.

If biofilm elimination; absence or reduced biofilm structures as observed microscopically, and reduced wound size was reported, we included it as the primary outcome, while if complete wound closure; healed or unhealed or reduced signs of infection; erythema, edema, warmth, pain, and dysfunction were reported, we included them as the secondary outcomes. We included clinical studies conducted in all wound care settings, including in-patient and ambulatory facilities. Non-clinical studies (e.g., in vitro, product development involving subjects without wounds) were excluded.

### 5.2 Participants

Included participants were: all types of wounds (e.g., acute and chronic wounds, such as diabetic foot ulcers, venous ulcers, surgical site infections, etc.) or clinical studies and trials (e.g., randomized controlled trials, non-controlled interventional studies, observational studies involving human subjects). Participants were excluded if the articles were published in languages other than English or studies not related to wound biofilms.

### 5.3 Interventions

Studies were eligible if they evaluated the use of wound dressing or ointment to prevent, inhibit, or eliminate wound biofilms.

### 5.4 Comparators

We included: any topical wound interventions (e.g., topical agents or wound dressings, including normal saline, iodine, honey, etc.) deemed as the standard of care or among the products.

### 5.5 Outcomes

Studies with the following primary outcomes were included: biofilm elimination, absence or reduced biofilm structures observed microscopically, or reduced wound size. Secondary outcomes were: complete wound closure: healed or unhealed, or infections or signs of infection: erythema, edema, warmth, pain, and dysfunction.

### 5.6 Setting

Studies were conducted in the following settings: all wound care settings, including in-patient and ambulatory facilities.

### 5.7 Study design

We included: randomized controlled trials, observational studies (all types), cohort (longitudinal) studies, cohort studies, or case-control studies. Only randomized controlled trials were included in the meta-analysis.

### 5.8 Search strategy

We included keywords (Appendix 1) in the search string MeSH or other subject terms, search filters, and text words. AA, a wound care expert, designed the search. The search was created with The Systematic Review Accelerator (https://pubmed.ncbi.nlm.nih.gov/32004673/).

Searches were done in PubMed, ClinicalTrials.gov, and Google Scholar with predefined keywords in MeSH terms. The dates searched were inception to 19th August 2022 (see Appendix 1). We restricted our initial search to exclude certain publication types.

Conference abstracts, theses, articles in press, books or book chapters, and reviews did not appear in our search results. Only articles published in English were included. We manually checked the included studies’ reference lists, performed a backward citation analysis, and had meta-analysis studies.

### 5.9 Study screening and selection

#### 5.9.1 Screening

Screening by title and abstract was conducted by AA and KRA independently. After identification and abstract screening, full texts were retrieved for the remaining articles. Two authors (AA and RAP) reviewed the full texts against the inclusion criteria. Discrepancies were resolved by consensus. Figure 1 shows the PRISMA flow diagram for the selection process.

**Figure 1.**
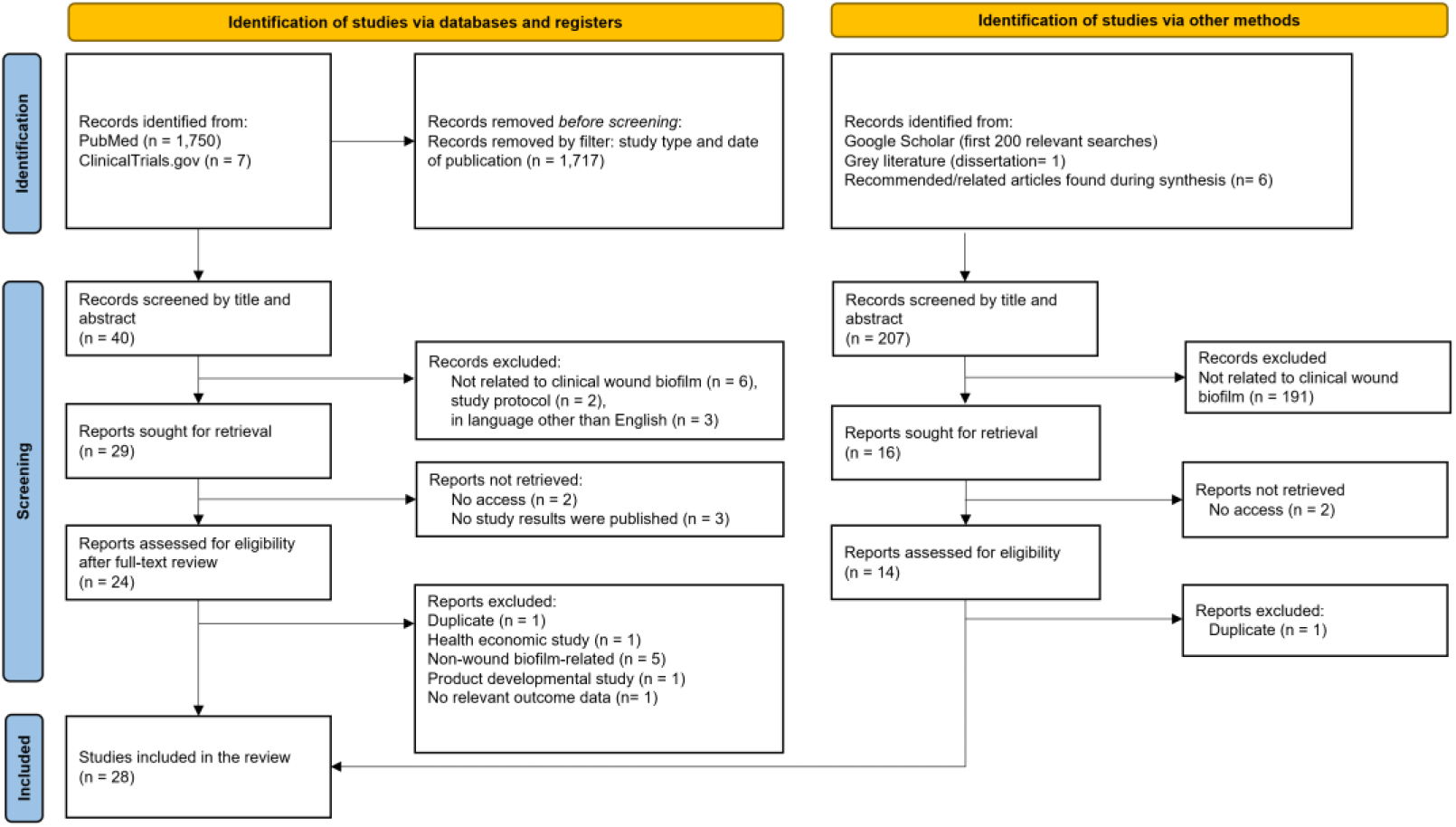
PRISMA Flow Diagram.

#### 5.9.2 Data extraction

We used a data extraction form for study characteristics and outcome data, piloted in Two studies in the review. Two study’s authors (AA & RAP) extracted the following data from included studies:

- types: randomized controlled trials, observational studies (all types), cohort (longitudinal) studies, cohort studies, case-control studies
- methods: study authors, year, study design, duration of follow-up, setting
- participants: number of participants, wound type(s)
- interventions and comparators: type of intervention, method of delivery, how the intervention was provided, frequency, type of comparator, no treatment, placebo, usual care
- outcomes: biofilm elimination; absence or reduced biofilm structures as observed microscopically (primary outcomes) or reduced wound size, complete wound closure; healed or unhealed or infections/signs of infection: erythema, edema, warmth, pain, and dysfunction; or bacterial load reduction (secondary outcomes). Table 1 describes the operational definition for each outcome.

**Table 1.**
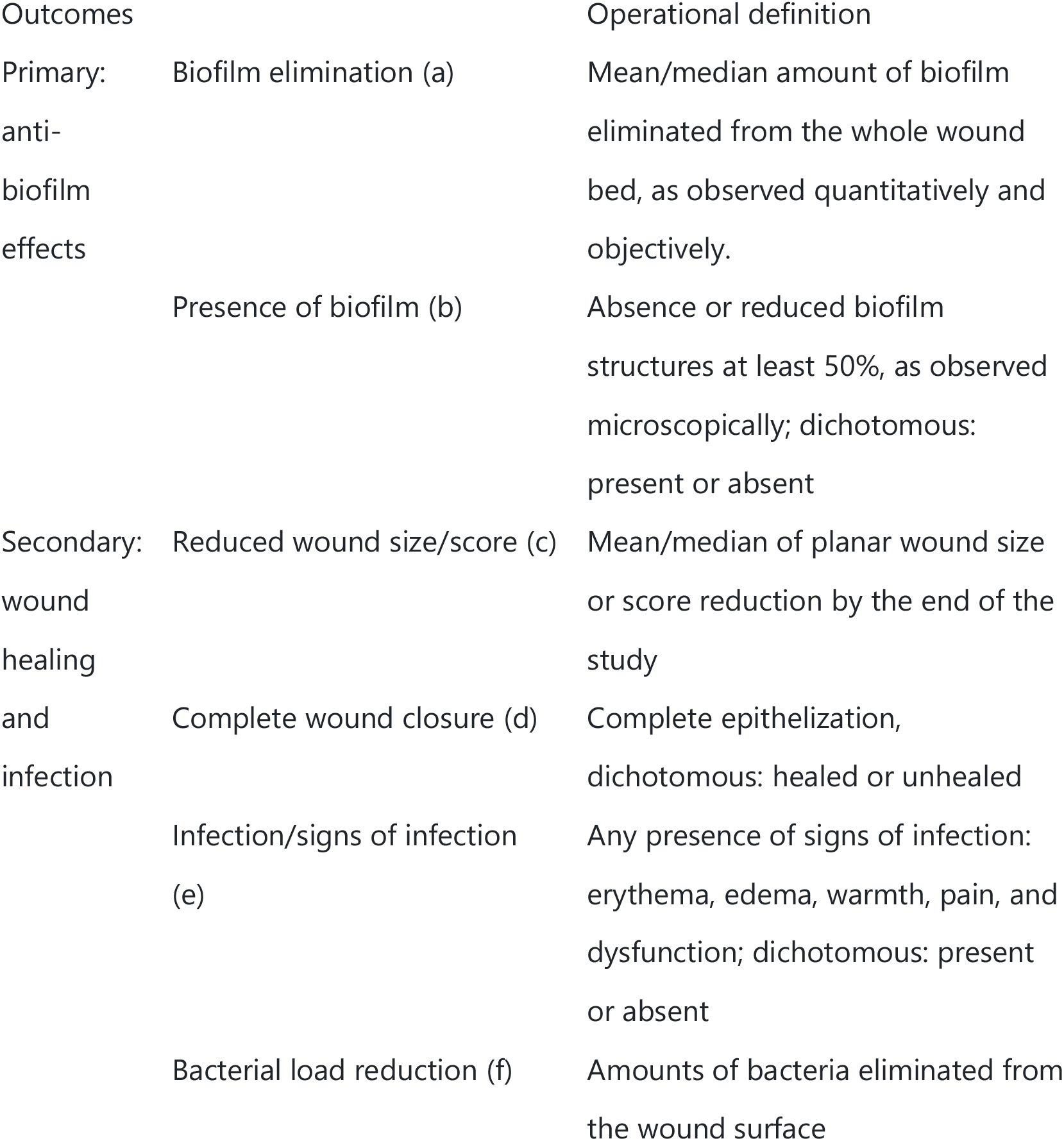
Operational definitions of the outcomes.

#### 5.9.3 Risk of Bias Assessment

Two review authors independently assessed the risk of bias for each study using the Cochrane Risk of Bias 2 tool.

### 5.10 Data analysis

The treatment/intervention effect was measured in Comprehensive Meta-Analysis v3.7. The treatment/intervention effect was calculated using log odds ratios (LOD) for dichotomous outcomes. We undertook meta-analyses when at least two studies or comparisons reported the same outcome. We used a random effects model. The unit of analysis was individual patients. We did not contact investigators or study sponsors to provide missing data.

We used the τ^2^ statistic to measure heterogeneity among the included trials. The publication bias / small studies effect was assessed using a Funnel plot.

We did not perform a subgroup analysis. Planned sensitivity analyses were: wounds receiving treatment for at least four weeks.

## 6. Results

A total of 1,964 records were identified by the systematic search, including 207 relevant searches in Google Scholar, grey literature, and articles found via reference tracing. Of those, 1,935 were excluded after the title and abstract screening and assessment of the eligibility criteria, as shown in Figure 1.

Of 28 articles, there are 13 RCTs (yielding 1,058 subjects), 3 case series, 8 interventional studies without controls, 3 observational studies, and 1 proof-of-concept interventional study. These studies include types of wound post-surgical^6,7^, venous^8–11^, diabetic foot ulcers (DFUs)^12–15^, leg ulcers^9,16–18^, pressure ulcers^19–21^, burns^22^, and unspecified chronic and acute ulcers^10,23,32,24–31^.

Of the RCTs, types of topical treatment sought for its anti-biofilm properties include 4 studies that used wound irrigation solution such as poly-hexamethylene biguanide (PHMB)^7,8,33^ and sodium hypochlorite^16^, 3 studies that used antibiotics^6,23,24^, 2 studies that used silver-based products^9,10^, 4 studies that used gel-type ointments^22,24^ and Iodosorb® (cadexomer iodine, Smith and Nephew, USA)^9,12^, 1 study that used negative pressure wound therapy with instillation^16^, and 2 studies that used the visual tool-guided wound cleansing^13,14^. The summary of each eligible article is shown in Table 2.

**Table 2.**
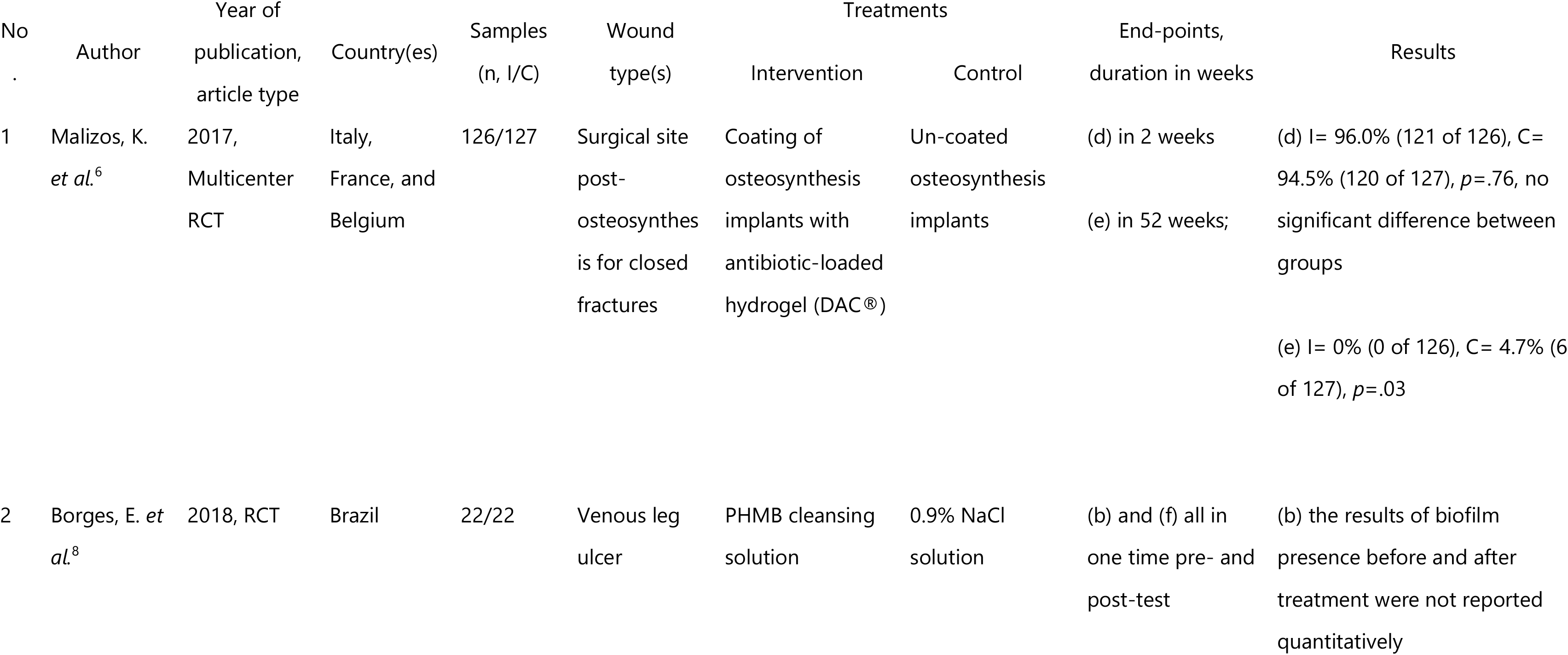
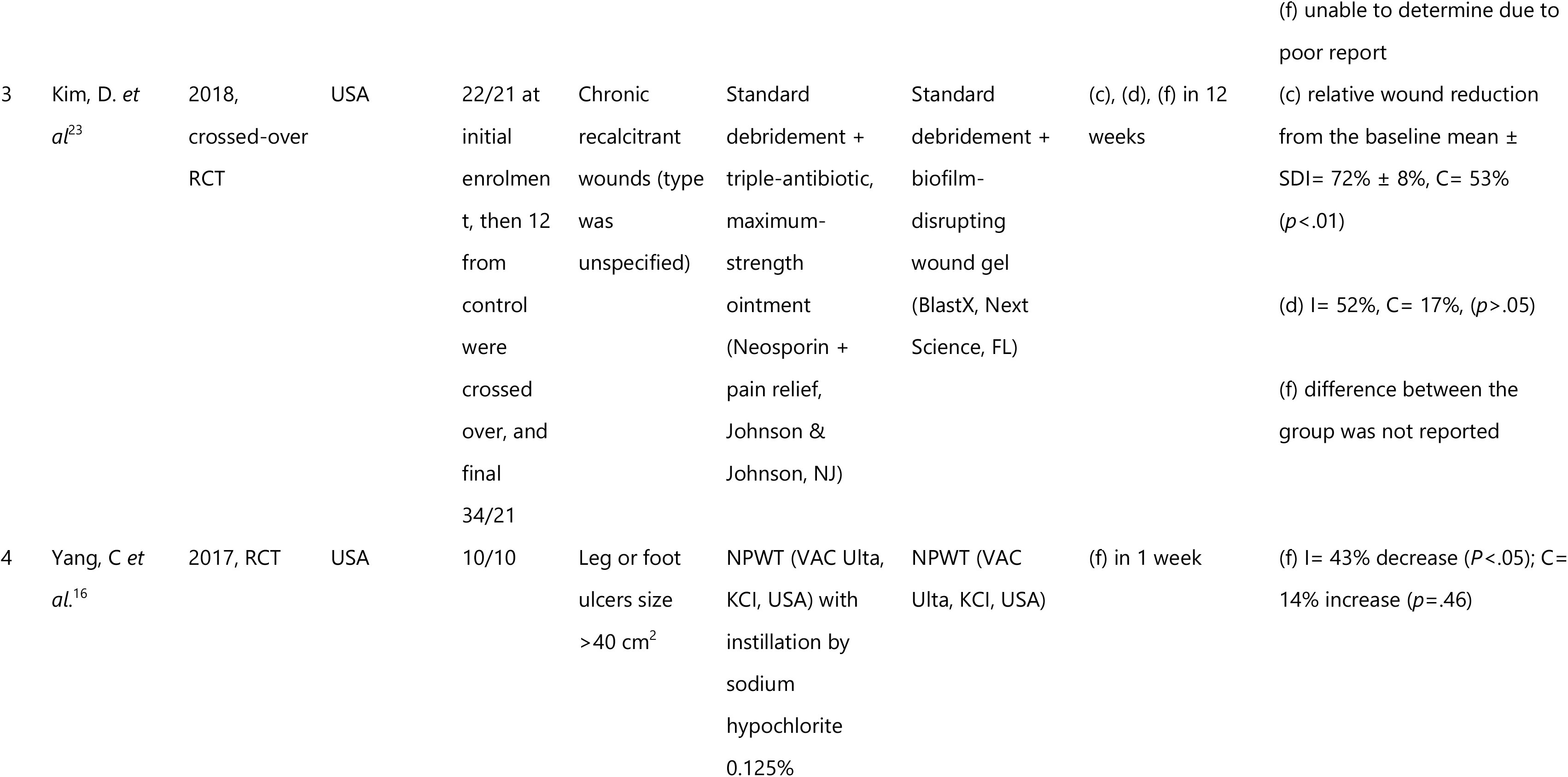
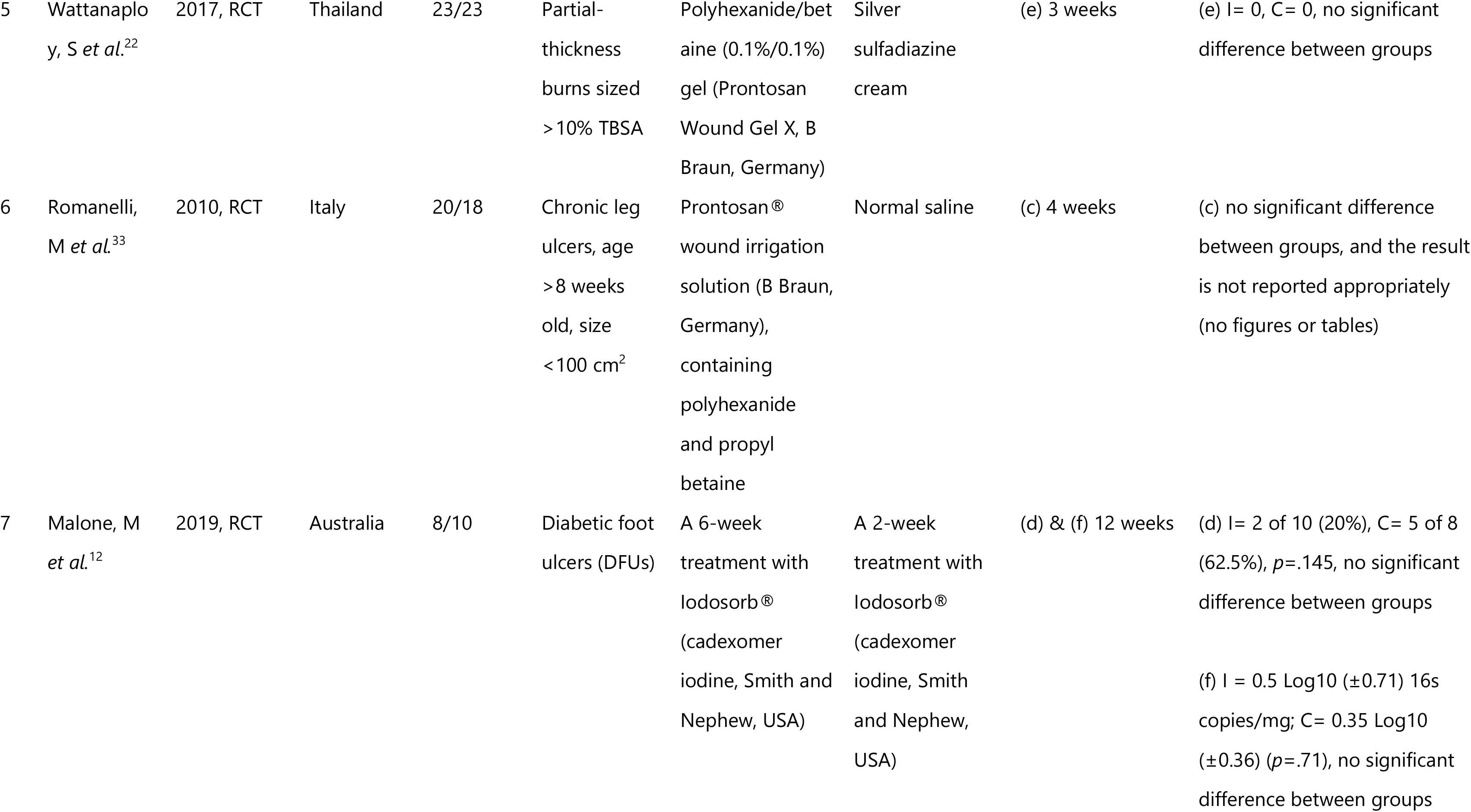
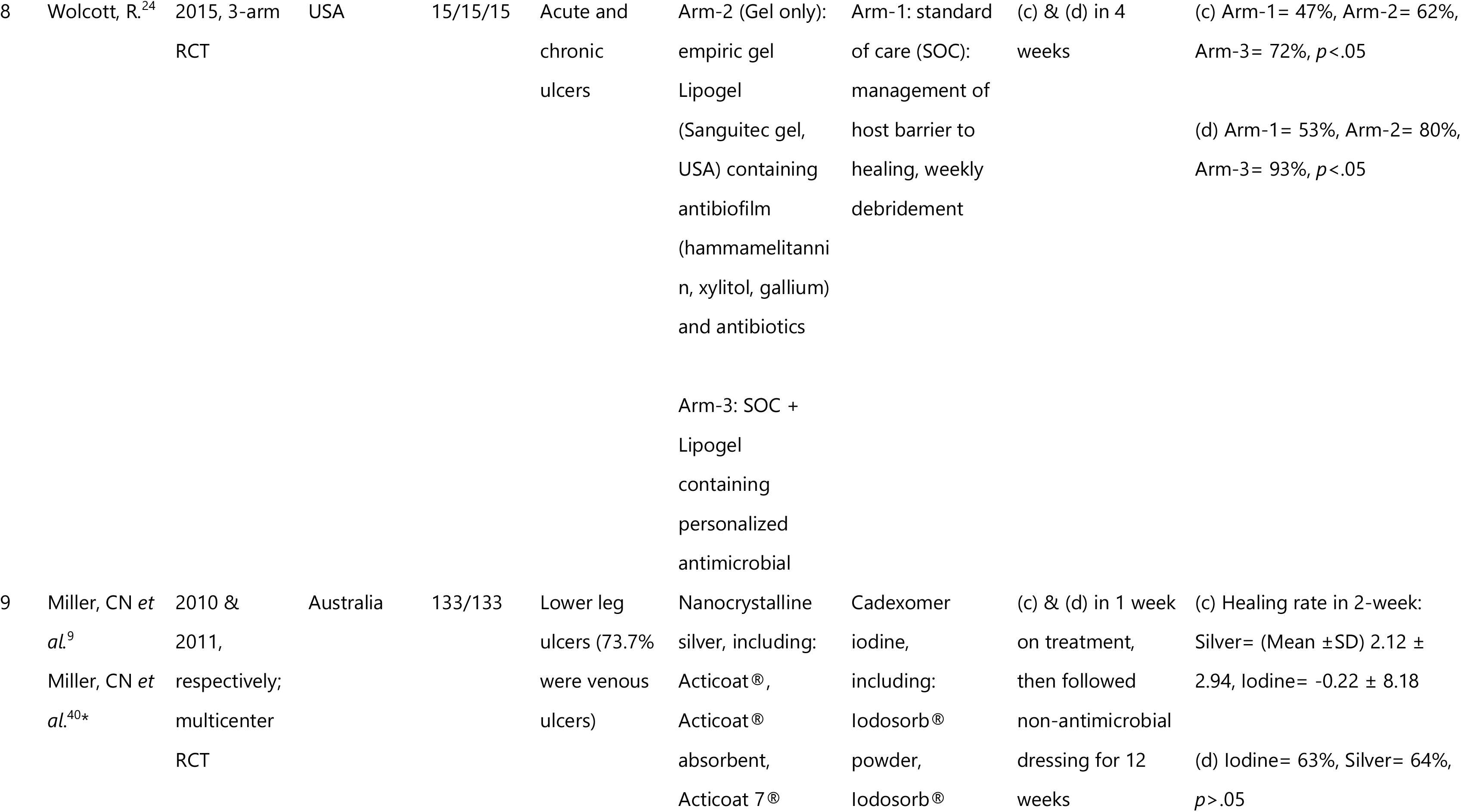
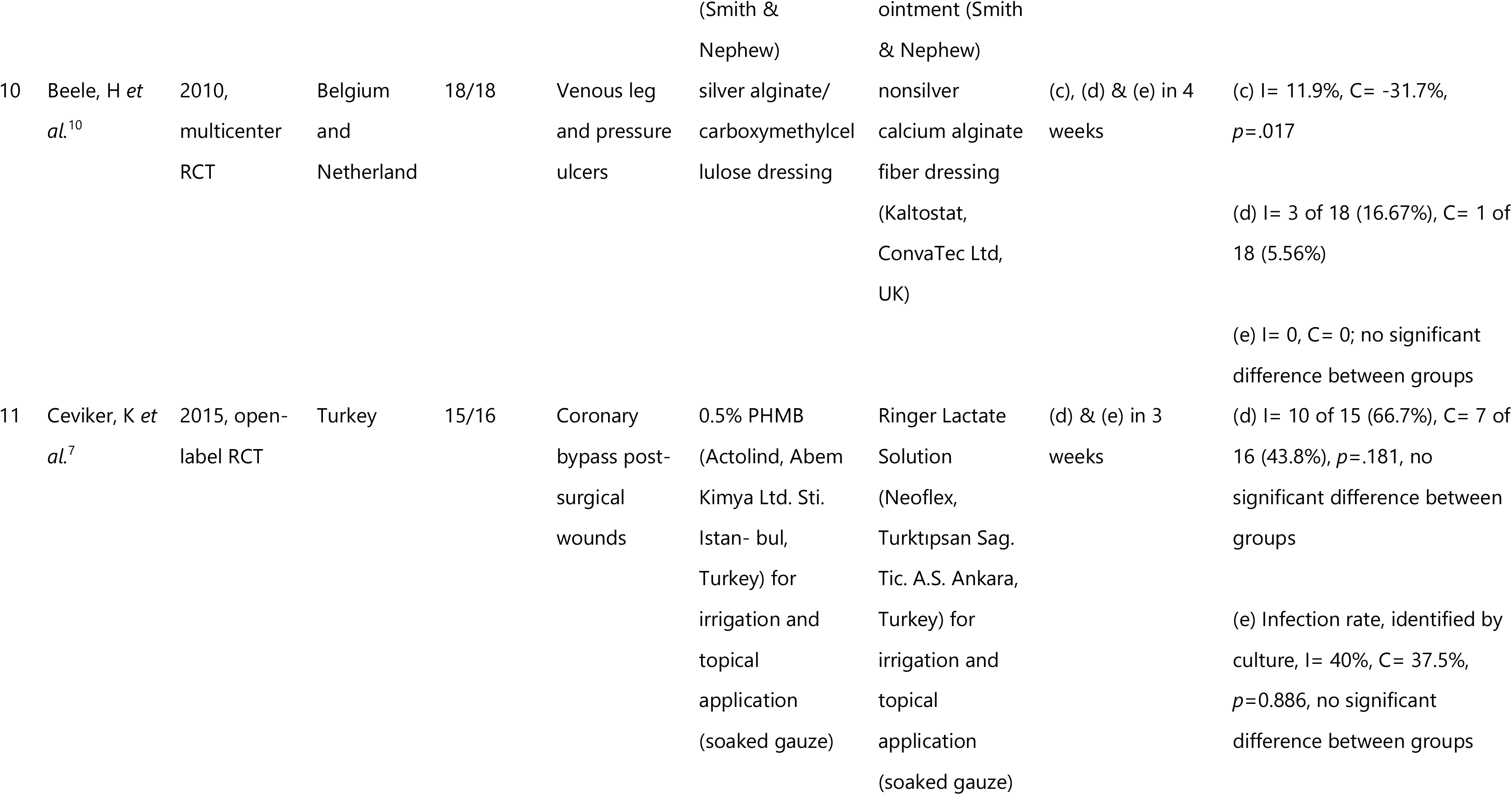
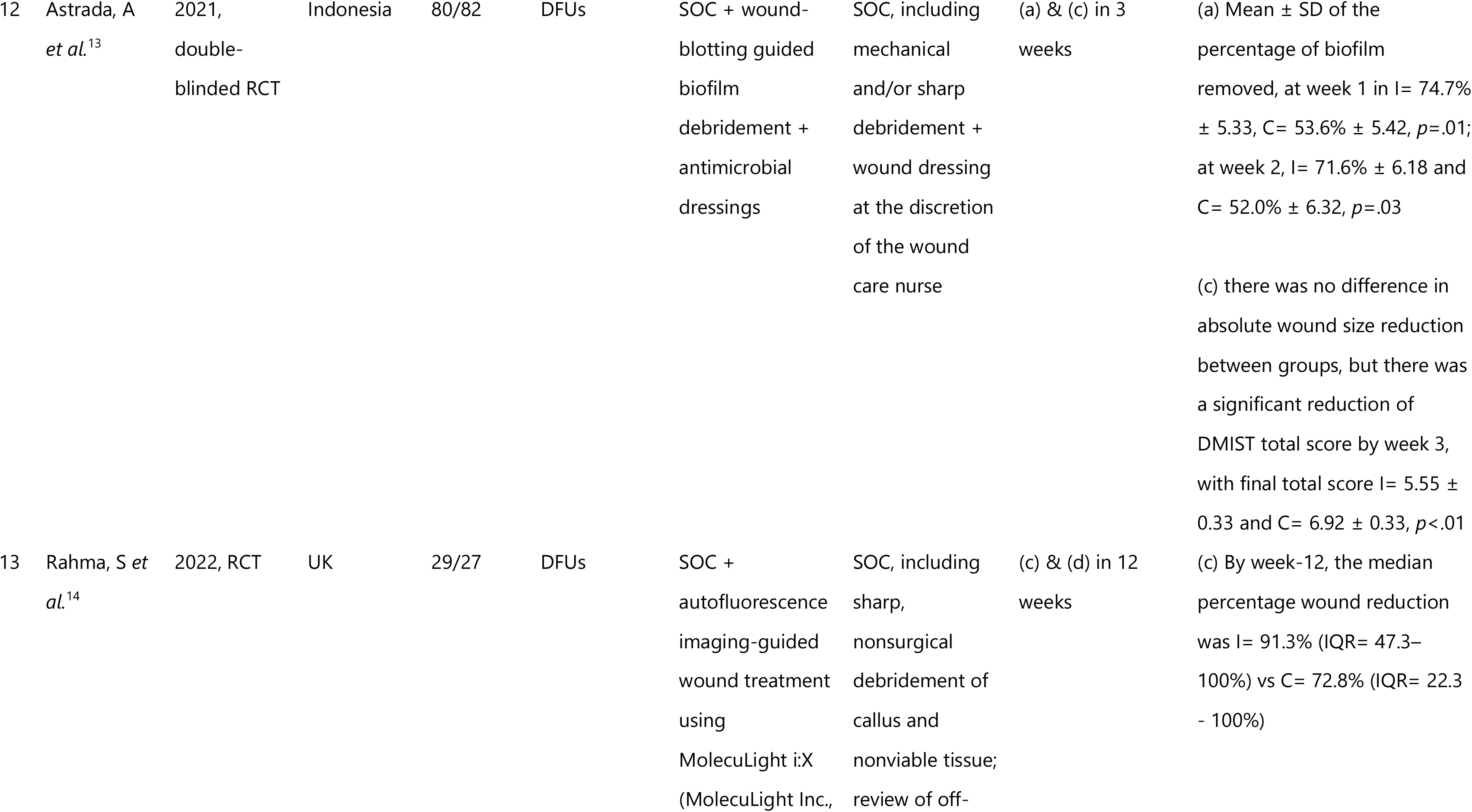
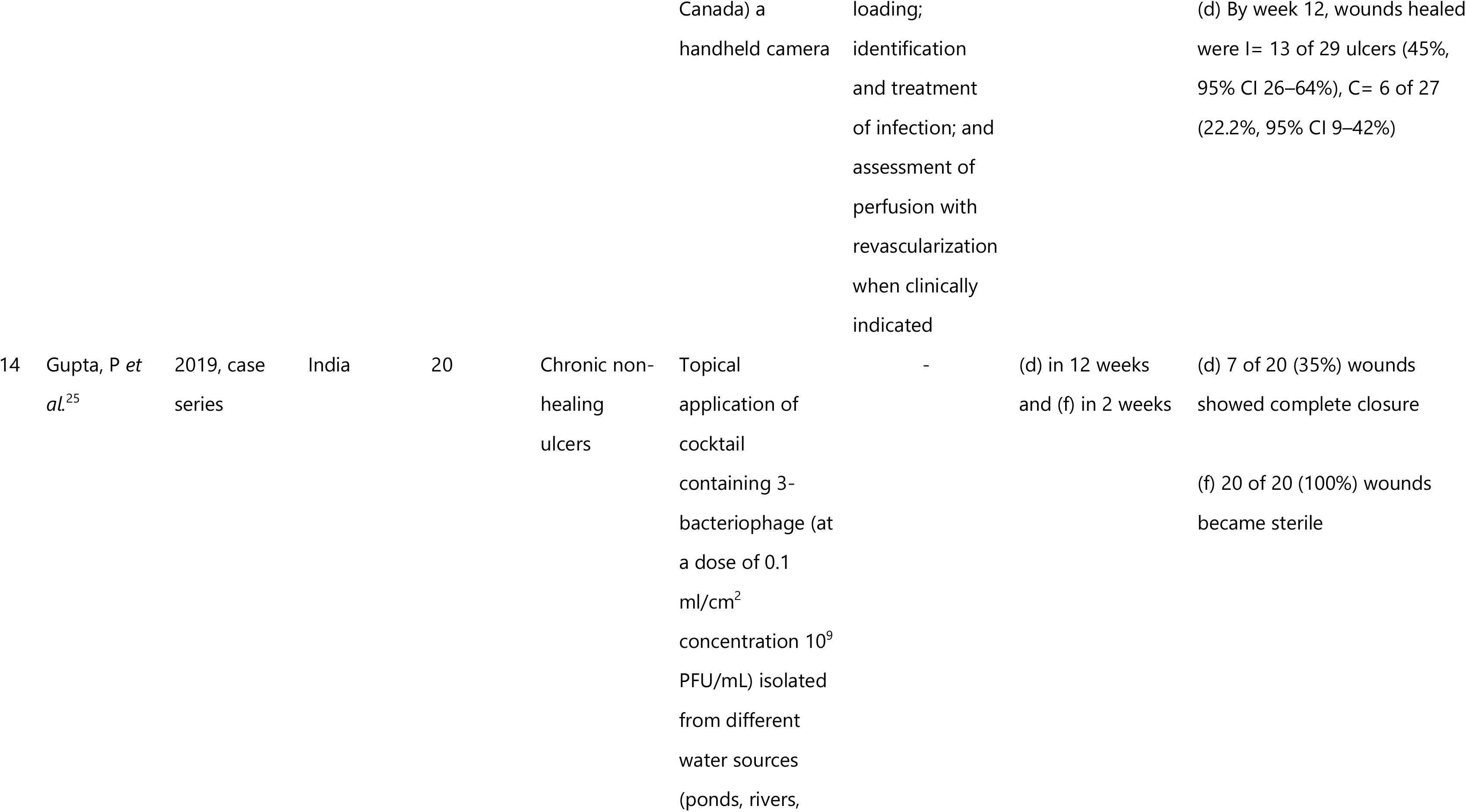
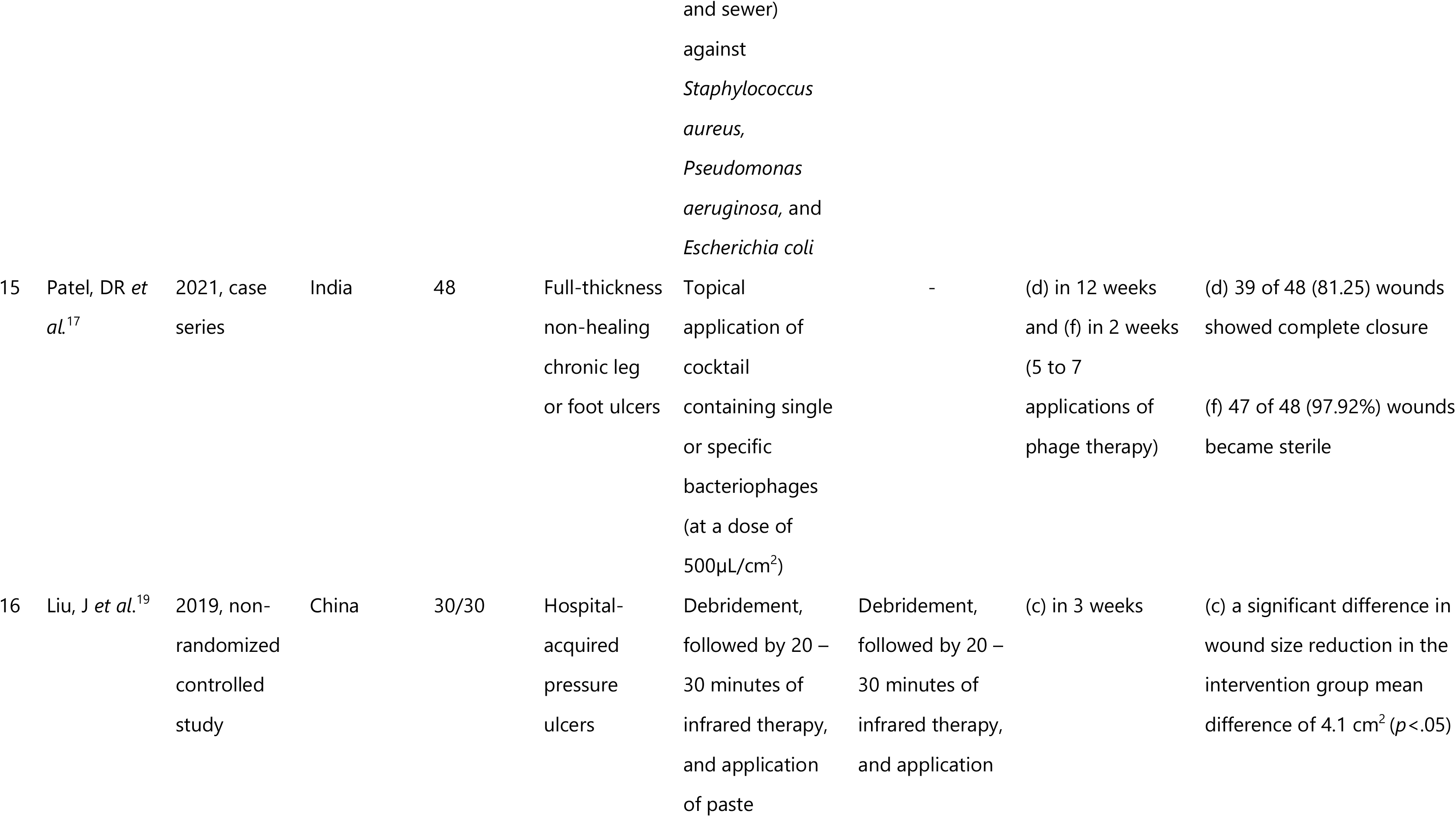
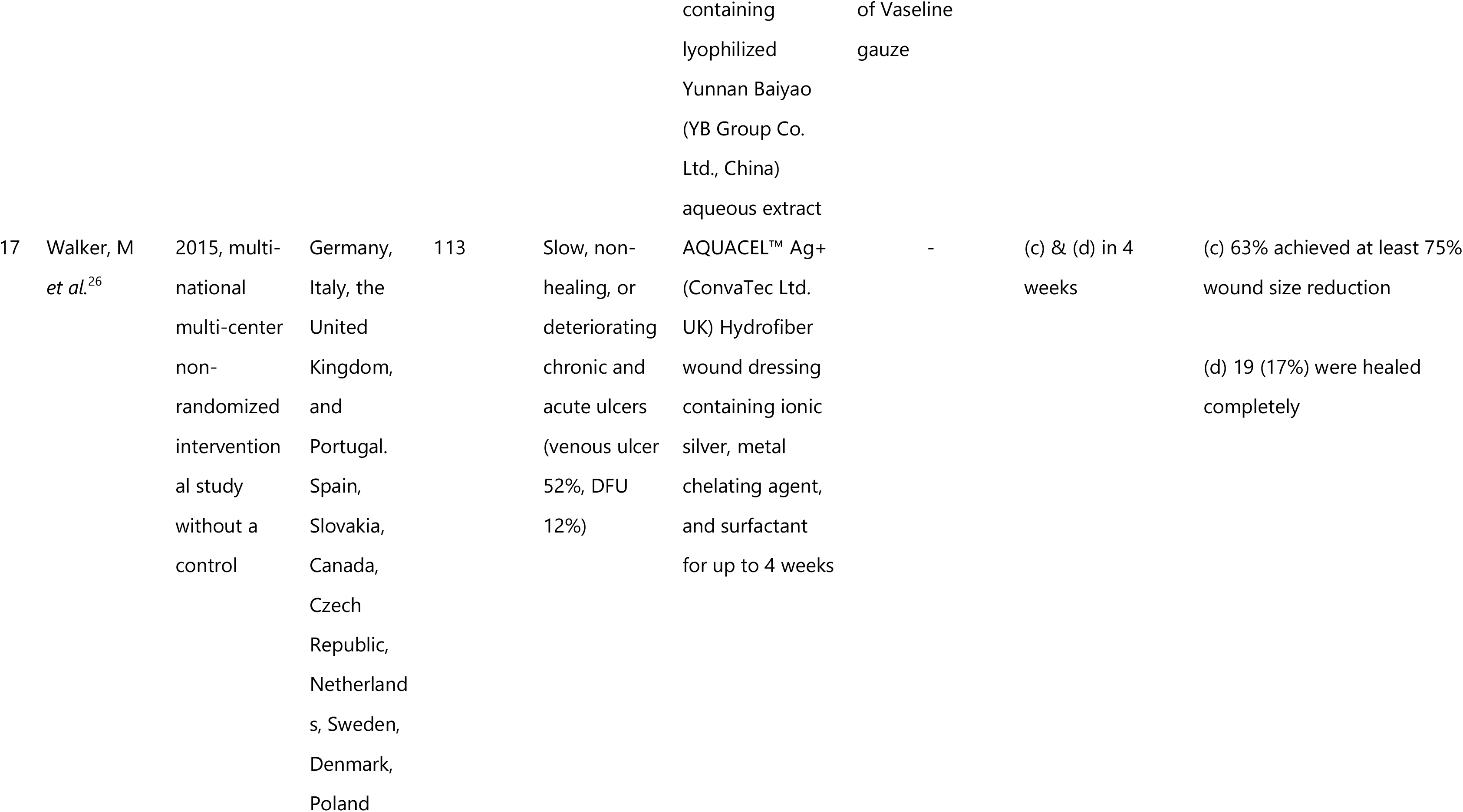
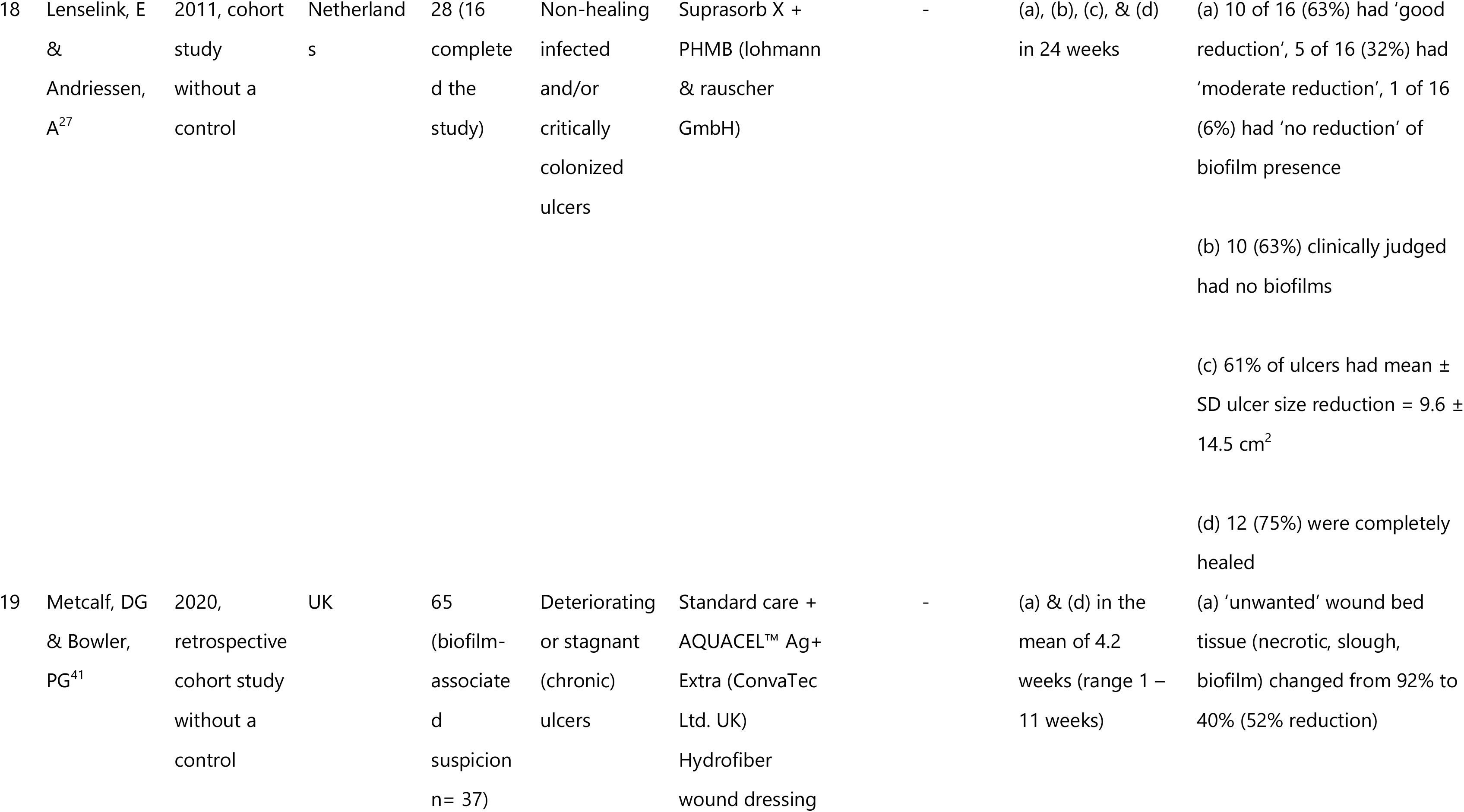
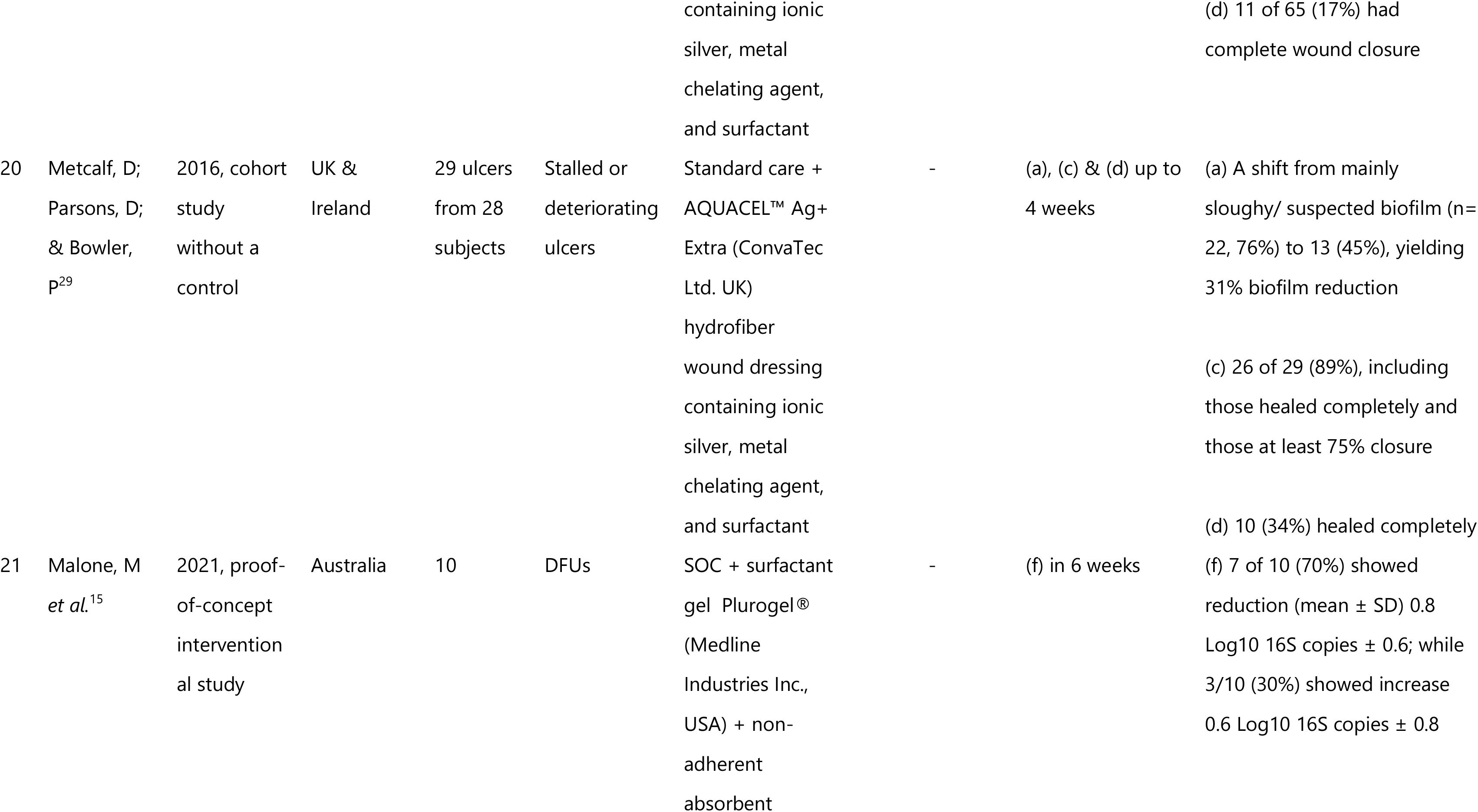
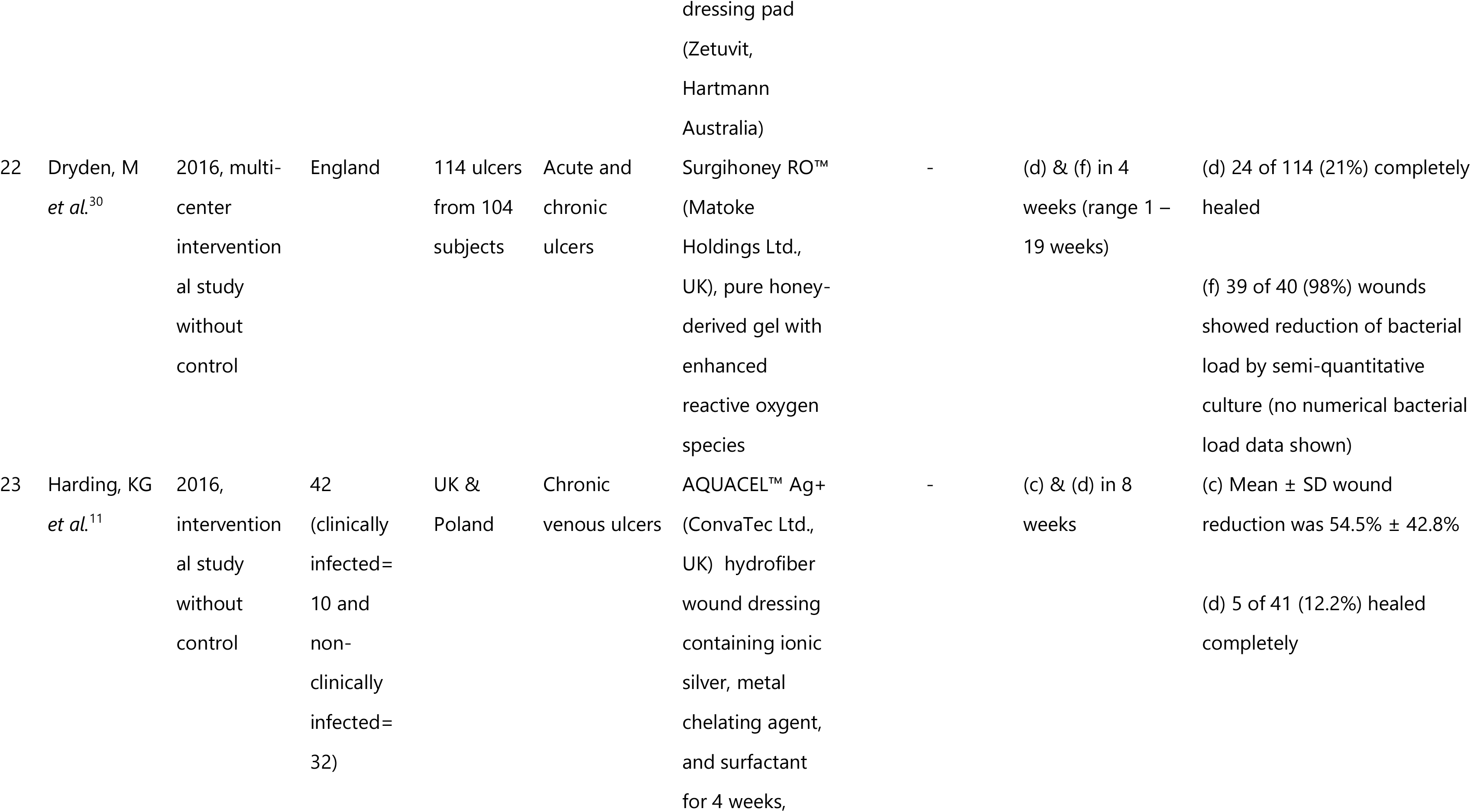
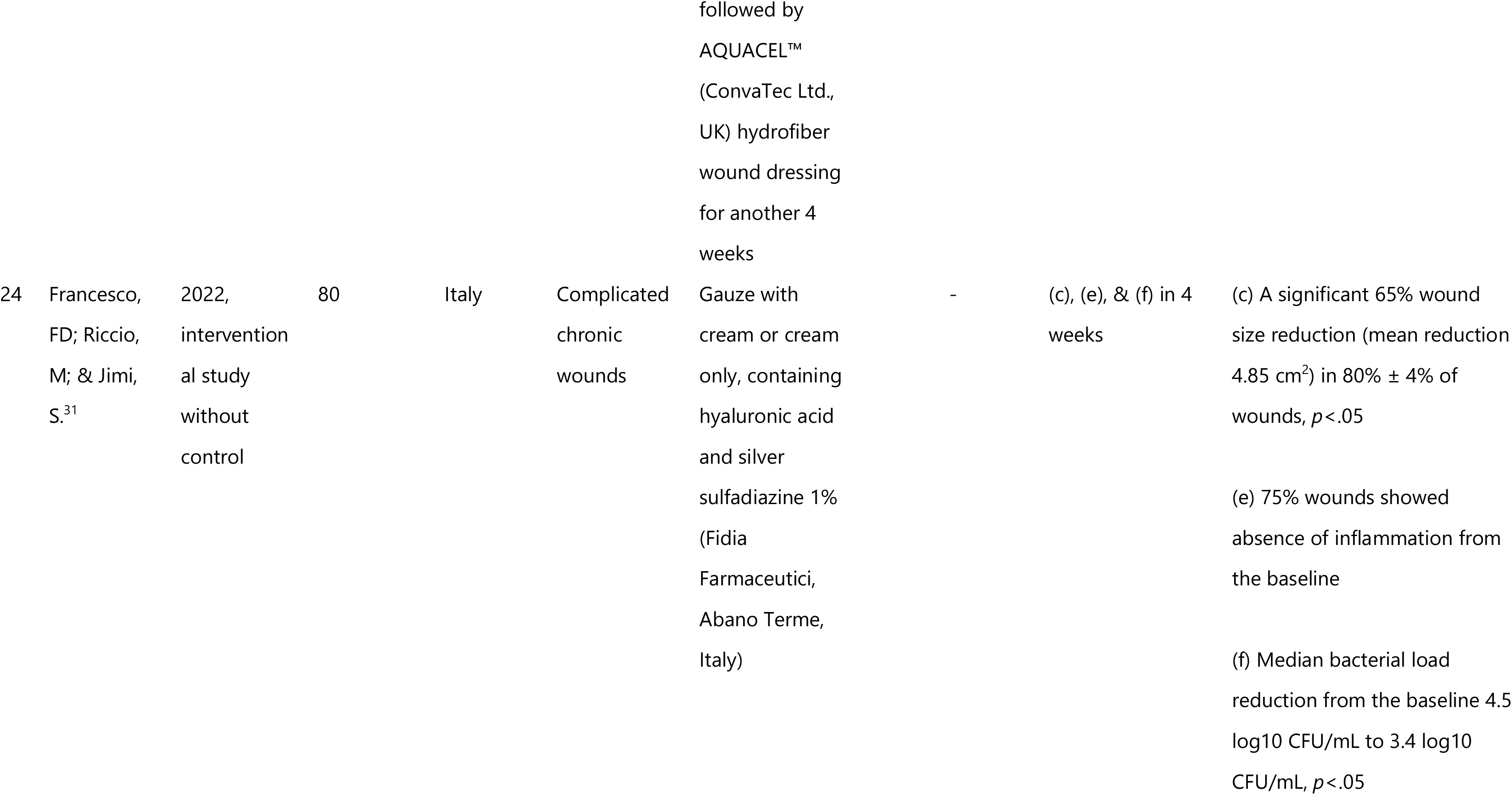
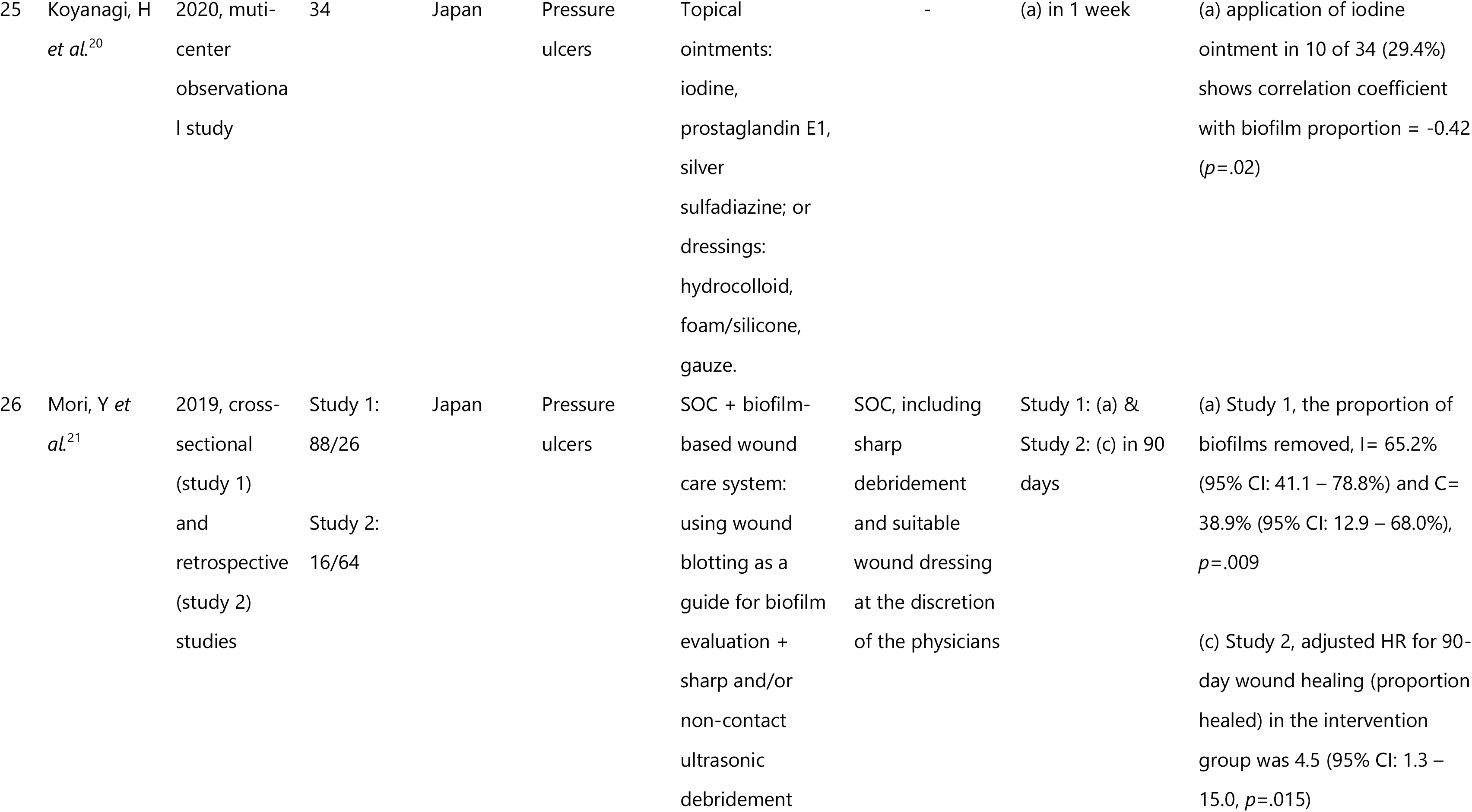
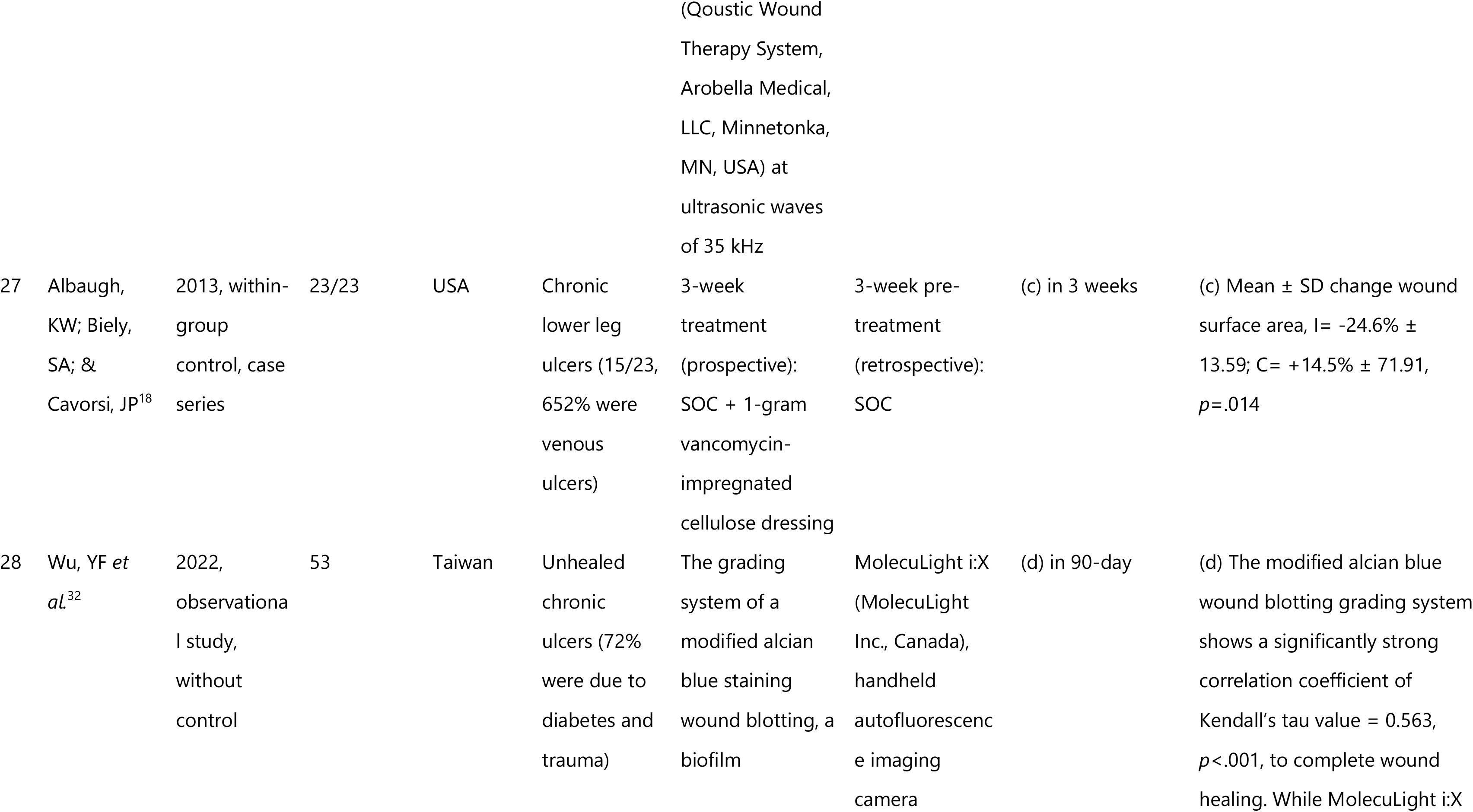
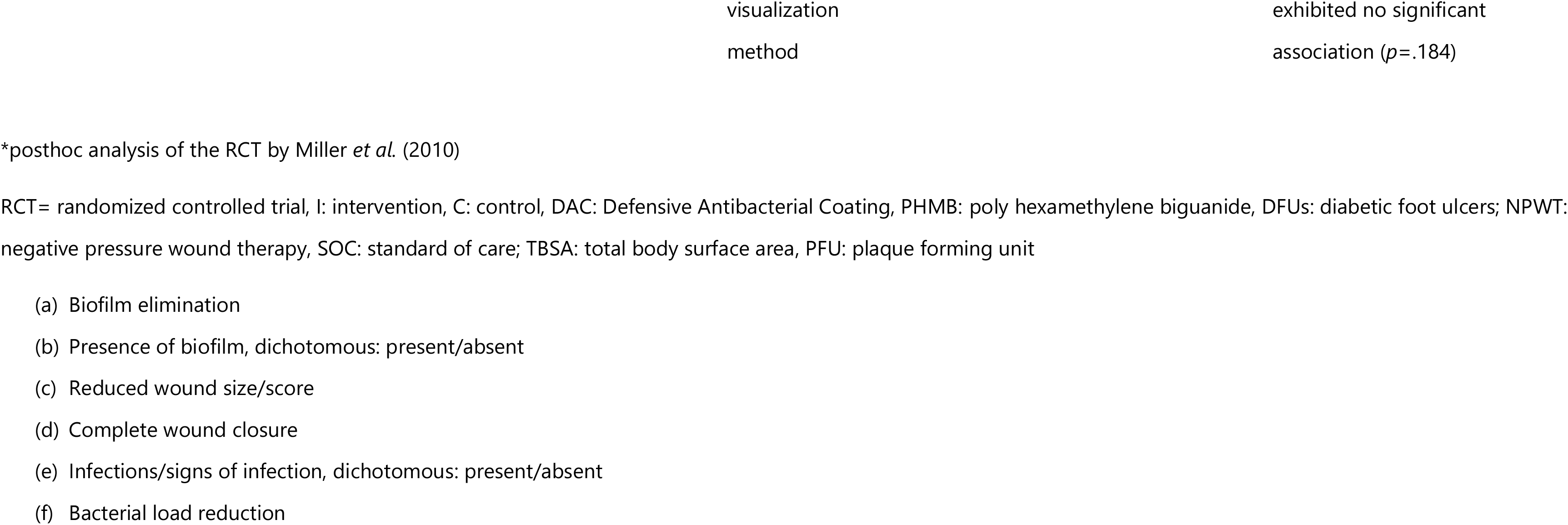
Summary of included studies.

### 6.1 Assessment of the Risk of Bias of the Included Studies

The risk of bias of all and individual RCTs is shown in Figures 2 and 3, respectively. All of the RCTs suggest a high risk (70%) of bias (Figure 2), primarily due to the detection bias and randomization process (selection bias) (Figure 3). Studies by Borges E *et al.* (2018) and Romanelli M *et al.* (2010) show a high risk of bias in all of the domains (Figure 3).

**Figure 2.**
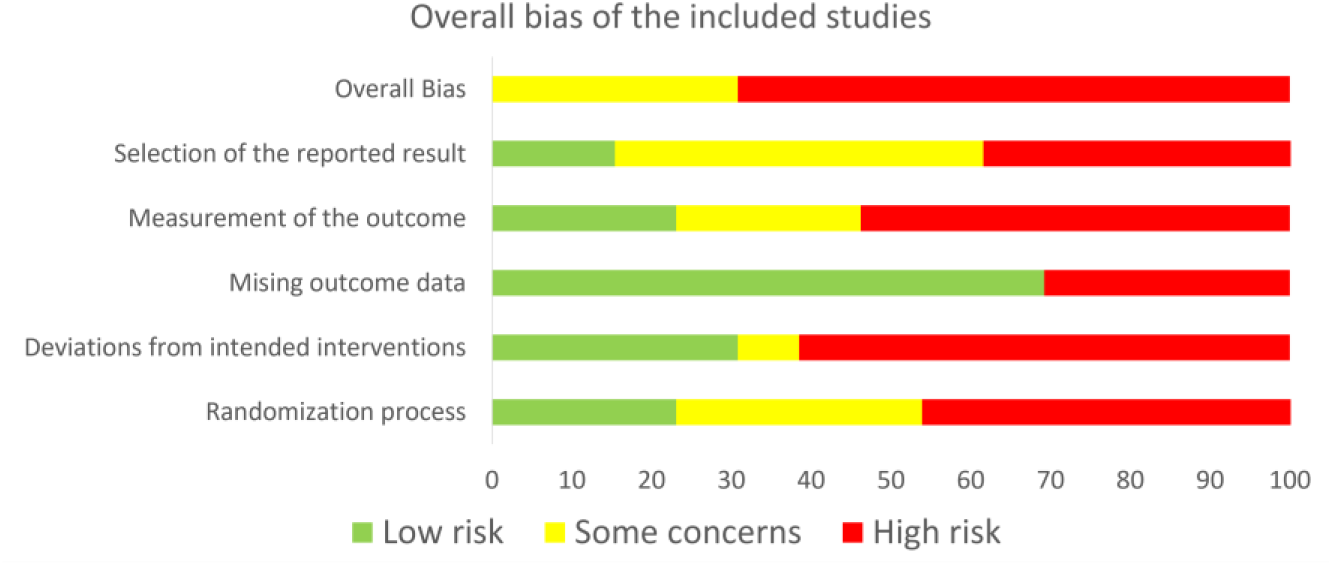
Risk of bias of all included RCTs.

**Figure 3.**
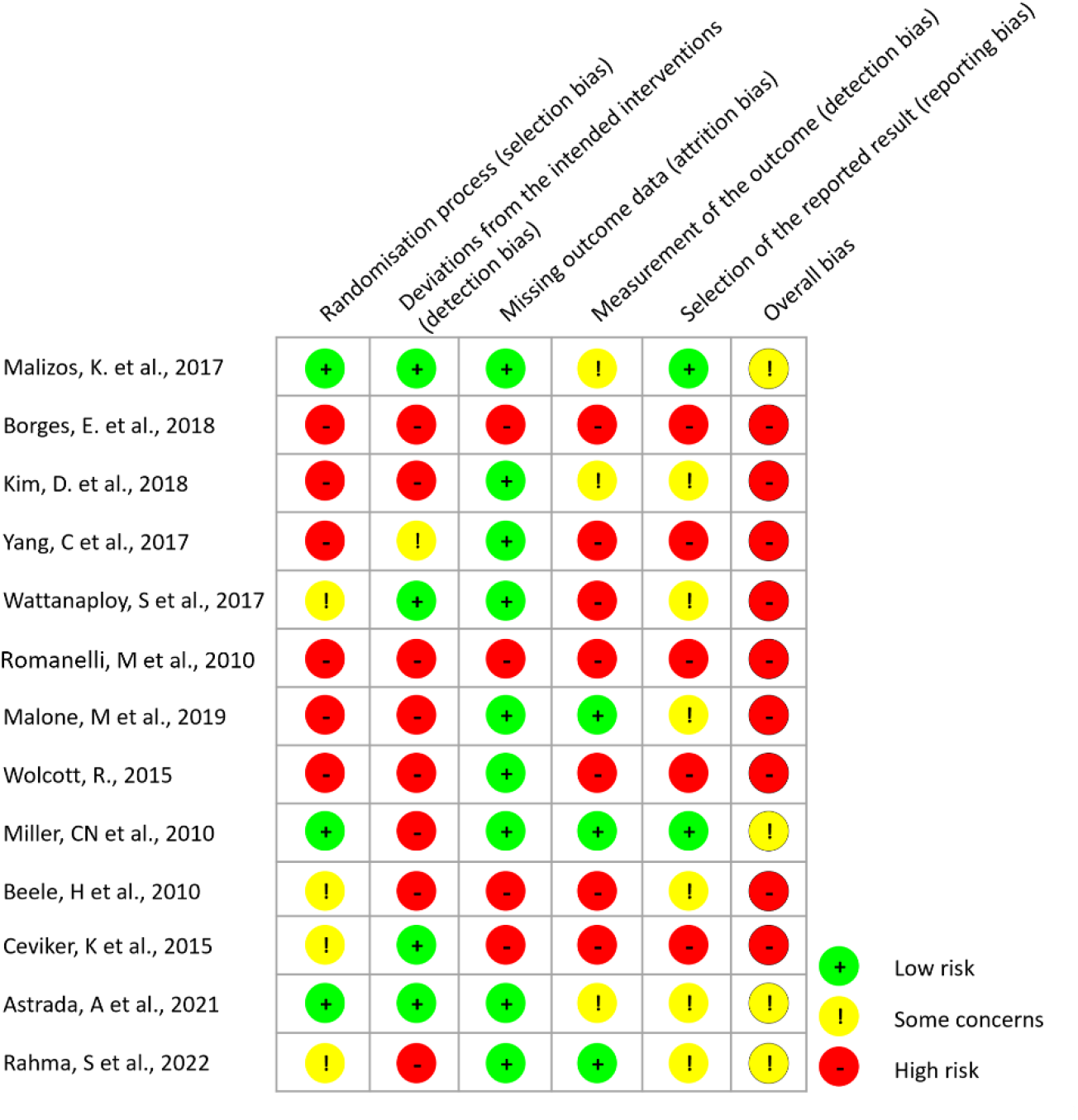
Risk of bias of individual RCTs.

### 6.2 Anti-Biofilm Effects of the Treatments

As the first primary outcome, the biofilm elimination is reported in one RCT by Astrada (2021)^13^; three interventional studies without controls by Lenselink *et al.* (2011)^27^ and Metcalf (2016 & 2020)^28,29^; and two observational studies by Mori *et al.* (2019)^21^ and Koyanagi *et al.* (2021)^20^ (Table 2). The meta-analysis was not feasible since only one study was found for each primary outcome.

Astrada *et al.* (2021) conducted a double-blinded randomized controlled trial in Indonesia. Patients with DFUs were treated with standard of care (SOC) along with wound-blotting guided biofilm debridement and antimicrobial dressings. The study demonstrated significant anti-biofilm effects, with the intervention group showing a mean ± SD percentage of biofilm removed of 74.7% ± 5.33 at week 1, compared to 53.6% ± 5.42 in the control group (p = 0.01). By week 3, a significant reduction in the DMIST total score was observed in the intervention group (5.55 ± 0.33) compared to the control group (6.92 ± 0.33, p < 0.01), indicating improved wound healing outcomes.

In the study by Lenselink *et al.* (2011), the patients were treated with Suprasorb X dressings containing PHMB. The study demonstrated that 63% of the patients showed “good reduction,” and 32% showed “moderate reduction” in biofilm presence.

Additionally, two studies by Metcalf *et al.* (2016 & 2020) investigated the effectiveness of AQUACEL™ Ag^+^ (ConvaTec Ltd. UK), a hydrofiber wound dressing containing ionic silver, metal chelating agent, and surfactant in addition to SOC on stagnant or deteriorating chronic ulcers in 4 weeks of intervention shows. In their year 2020’s study, they found that the ‘unwanted’ wound bed tissue, assumed as harboring biofilm, changed from 92% to 40% (52% reduction). While in their 2016’s study, they demonstrated a 31% reduction in the wounds showing biofilm presence.

The observational studies by Mori et al. (2019) and Koyanagi *et al*. (2021) also highlighted the use of the wound-blotting method to evaluate biofilm elimination. In a cross-sectional and retrospective study conducted in Japan by Mori et al. (2019), two studies evaluated the effectiveness of a biofilm-based wound care system in pressure ulcers. The intervention group showed significant anti-biofilm effects, with a higher proportion of biofilm removal (65.2% vs. 38.9%, p = 0.009) and an increased likelihood of 90-day wound healing (adjusted HR: 4.5, 95% CI: 1.3 −15.0, p = 0.015) compared to the control group.

The second primary outcome, biofilm presence, is reported only in one RCT by Borges *et al.* (2018)^8^ and one interventional studies without control by Lenselink *et al.* (2011)^27^ (Table 2). Borges *et al.* (2018) conducted an RCT in Brazil involving patients with venous leg ulcers. The intervention group received treatment with a PHMB cleansing solution, while the control group received a 0.9% NaCl solution. However, the study did not provide quantitative statistical findings on biofilm presence before and after treatment. Thus, the results regarding the effect of the intervention on biofilm presence are inconclusive.

Lenselink *et al.* (2011)^27^, Metcalf *et al.* (2016 & 2020)^28,29^, and Borges *et al.* (2018)^8^ evaluated the primary outcomes only by surrogate or clinical cues such as the presence of necrotic tissue or visible gel-like materials found on wound surface (Table 2). Astrada *et al.* (2021), Mori *et al.* (2019)^21^, and Koyanagi *et al.* (2021)^20^ used the wound-blotting method to evaluate the primary outcomes (Table 2).

### 6.3 Wound Healing

For the secondary outcomes, the reduced wound size/score is reported in 6 RCTs^9,10,13,14,24,33^, in 6 case series or non-controlled interventional studies^11,19,26,27,31,34^, and 2 observational studies^18,35^. However, the meta-analysis for this outcome could not be performed due to the poor data report^33^, and data was ununifiable^9,10,13,14,24^.

An RCT conducted by Romanelli et al. (2010) in Italy assessed the efficacy of Prontosan® wound irrigation solution compared to normal saline in treating chronic leg ulcers. After a 4-week, no significant difference in wound size reduction was observed between the groups.

In a 3-arm RCT conducted by Wolcott *et al.* (2015) in the USA, the effectiveness of different treatments on acute and chronic ulcers was evaluated. Arm-1 represented the control group, receiving SOC management. Arm-2 received Lipogel (Sanguitec gel, USA) containing antibiofilm (hammamelitannin, xylitol, gallium) and antibiotics, and Arm-3 received SOC along with Lipogel containing personalized antimicrobial. The study included 15 subjects in each arm, and after four weeks of weekly debridement, Arm-3 demonstrated the most significant improvement in wound size reduction (72%) compared to Arm-1 (47%) and Arm-2 (62%), with statistical significance.

Two multicenter RCTs investigated silver-based products. Miller *et al.* (2010) conducted a multicenter RCT in Australia to compare the effects of nanocrystalline silver and cadexomer iodine treatments on lower leg ulcers, most of which were venous ulcers.

After two weeks of treatment with antimicrobial dressings, the mean wound size reduction was 2.12 ± 2.94 cm^2^ in the silver group and −0.22 ± 8.18 cm^2^ in the iodine group. In Belgium and the Netherlands, Beele et al. (2010) investigated the efficacy of silver alginate/carboxymethylcellulose dressing compared to non-silver calcium alginate fiber dressing in the venous leg and pressure ulcers. After a 4-week duration, the intervention group showed a wound size reduction of 11.9%, whereas the control group exhibited an increase of 31.7% (*p*<.05).

While in the same category of silver-based products, interventional studies without control by Walker *et al.* (2015), Metcalf *et al.* (2016), and Harding *et al.* (2016) investigated the effectiveness of AQUACEL™ Ag^+^ (ConvaTec Ltd. UK) in addition to SOC on stagnant or deteriorating chronic ulcers in 4 weeks of intervention. They found that the mean wound reduction ranged from 54% to 89%. Additionally, Francesco *et al.* (2022) investigated the use of cream containing SSD and hyaluronic acid in chronic wounds for four weeks. They found 65% wound size reduction (mean reduction 4.85 cm^2^) in 80% ± 4% of wounds, p<.05.

Two visual tool-guided RCTs were conducted by Astrada *et al.* (2021), and Rahma *et al.* (2022).^14^ Astrada *et al.* (2021)^13^ conducted a double-blinded RCT in Indonesia to evaluate the impact of wound-blotting guided biofilm debridement and antimicrobial dressings on DFUs compared to SOC. After a 3-week, there was no significant difference in absolute wound size reduction between the groups. However, the intervention group significantly reduced the DMIST^36^ total score by week 3 (p<.01), indicating improved wound characteristics. While Rahma *et al.* (2022) conducted an RCT in the UK to assess, the efficacy of autofluorescence imaging-guided wound treatment using MolecuLight i:X compared to SOC in DFUs. After 12 weeks, the intervention group exhibited a median percentage wound reduction of 91.3% (IQR=47.3–100%), while the control group had a median reduction of 72.8% (IQR=22.3-100%).

Antibiotic-based use is demonstrated in a within-group control case series conducted by Albaugh *et al.* (2013)^18^. They applied a 1-gram vancomycin-impregnated cellulose dressing on chronic wounds for three weeks. They showed mean ± SD = 24.6% ± 13.59 of wound size reduction in the intervention group, while in the control group, the size seems to increase by 14.5% ± 71.91 (*p*=.014).

Furthermore, a Chinese Medicine-based product by Liu *et al.* (2019) is presented in a non-randomized controlled study on 60 pressure ulcers, 30 each for intervention and control groups. They investigated the application of paste containing lyophilized Yunnan Baiyao (YB Group Co. Ltd., China) aqueous extract in addition to debridement and 20 – 30 minutes of infrared therapy. After three weeks, they found a significant difference in wound size reduction in the intervention group mean difference of 4.1 cm2.

While for the complete wound closure, it is reported in 8 RCTs^6,7,9,10,12,14,23,24^, in 8 case series or non-controlled interventional studies^11,17,25–28,30,34^, and 1 observational study^32^. Studies by Kim *et al.* (2018)^23^ and Miller *et al.* (2010)^9^ were not included in the analysis because of the unreported number of events of the wounds reaching closure.

The six RCTs included in the pooled analysis yielding n=439, including the studies by Malizos *et al.* (2017)^6^, Malone *et al.* (2019)^12^, Wolcott *et al.* (2015)^24^, Beele *et al.* (2010)^10^, Ceviker *et al.* (2015)^7^, and Rahma *et al.* (2022)^14^.

Two antibiotic-based studies investigated this outcome by Malizos *et al.* (2017) and Wolcott *et al.* (2015). Malizos *et al.* (2017) conducted a multi-center RCT across Italy, France, and Belgium, focusing on surgical site post-osteosynthesis for closed fractures. The intervention group (n=126) used the coating of osteosynthesis implants with antibiotic-loaded hydrogel (DAC®), while the control group (n=127) received un-coated osteosynthesis implants. After two weeks, wound closure in the intervention group was 96.0% (121 of 126), while in the control group was 94.5% (120 of 127), p=.76; no significant difference was observed between groups. On the other hand, a study by Wolcott et al. (2015) found that groups receiving Lipogel in combination with antibiotics and antibiofilm showed 80% to 93% of wounds achieved closure, while the control group only had 53%. However, there was no estimation of statistical difference provided.

The silver-based product study by Beele *et al.* (2010) also showed no significant difference between groups regarding wound closure.

Malone et al. (2019) conducted an RCT in Australia focusing on the effectiveness of a 6-week vs. 2-week treatment with Iodosorb® (cadexomer iodine, Smith and Nephew, USA) on DFUs. The study involved eight subjects in the intervention group and ten in the control group. After 12 weeks, the incidence of DFU complete closure was observed in 2 of 10 subjects (20%) in the intervention group, compared to 5 of 8 subjects (62.5%) in the control group. However, the difference between the groups was not statistically significant (p=.145).

Ceviker et al. (2015) conducted an open-label RCT focusing on coronary bypass post-surgical wounds in Turkey. The intervention group (n=15) received 0.5% PHMB (Actolind) for irrigation and topical application (soaked gauze), while the control group (n=16) received Ringer Lactate Solution (Neoflex) for irrigation and topical application (soaked gauze). After a 3-week, the complete wound closure was 66.7% (10 of 15) in the intervention group and 43.8% (7 of 16) in the control group, with significant differences between groups.

The visual-guided wound intervention study by Rahma *et al.* (2022) shows that by week 12, wounds receiving the autofluorescence imaging-guided wound treatment had the number of wounds healed 13 of 29 (45%, 95% CI 26–64%), while in the control group 6 of 27 (22.2%, 95% CI 9–42%). There was no statistical significance estimated.

Figure 4 shows a forest plot of the pooled random effects of 6 RCTs for this outcome with a log odds ratio (LOD) of 0.03 (95% CI: −1.02 – 1.09). The between-study heterogeneity variance was estimated at τ^2^=1.09 (SE: 1.10; Q=14.33, *p*=.014). The funnel plot of this outcome is shown in Figure 5.

**Figure 4.**
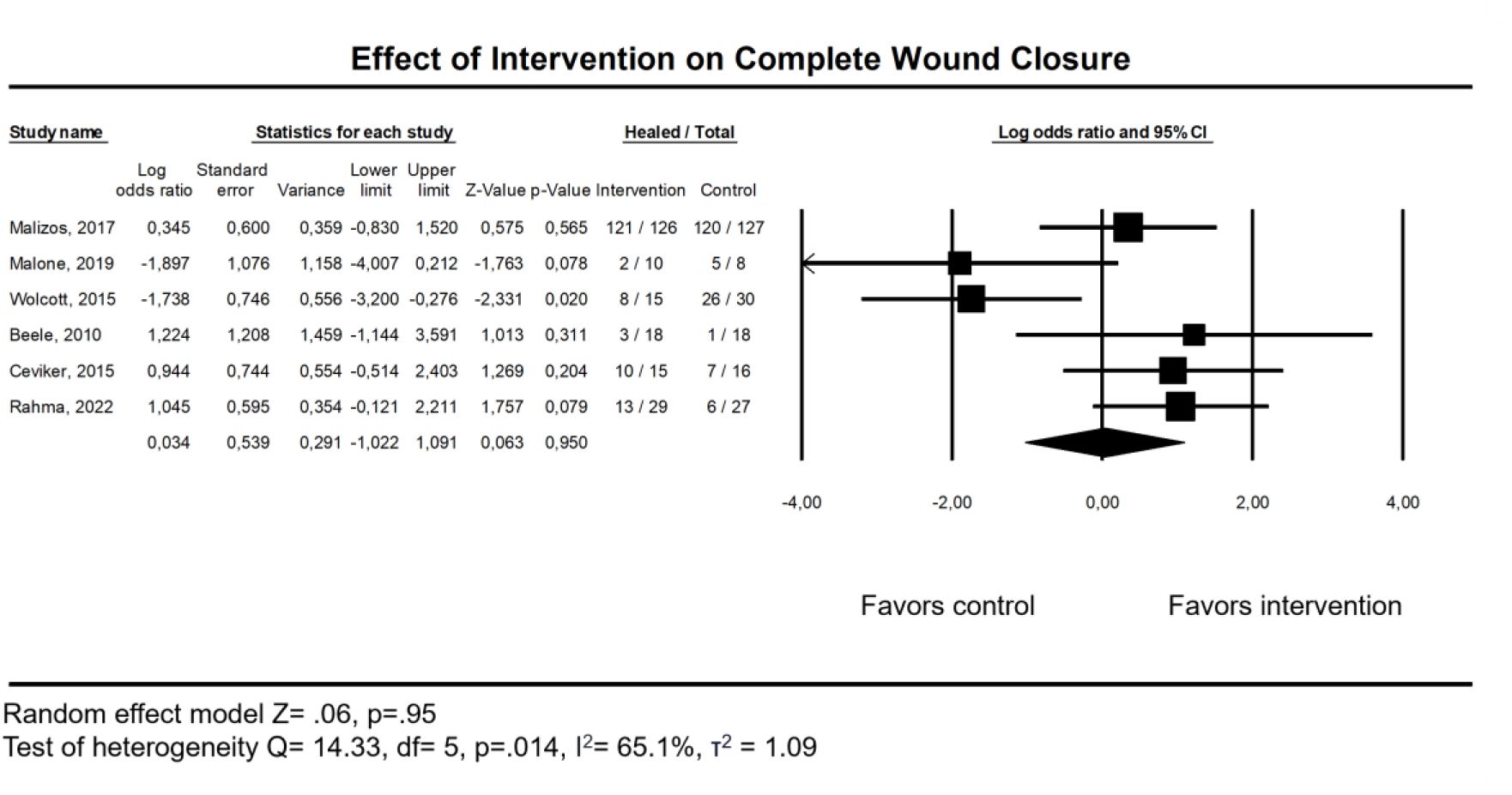
Forest plot of the effect of intervention on complete wound closure.

**Figure 5.**
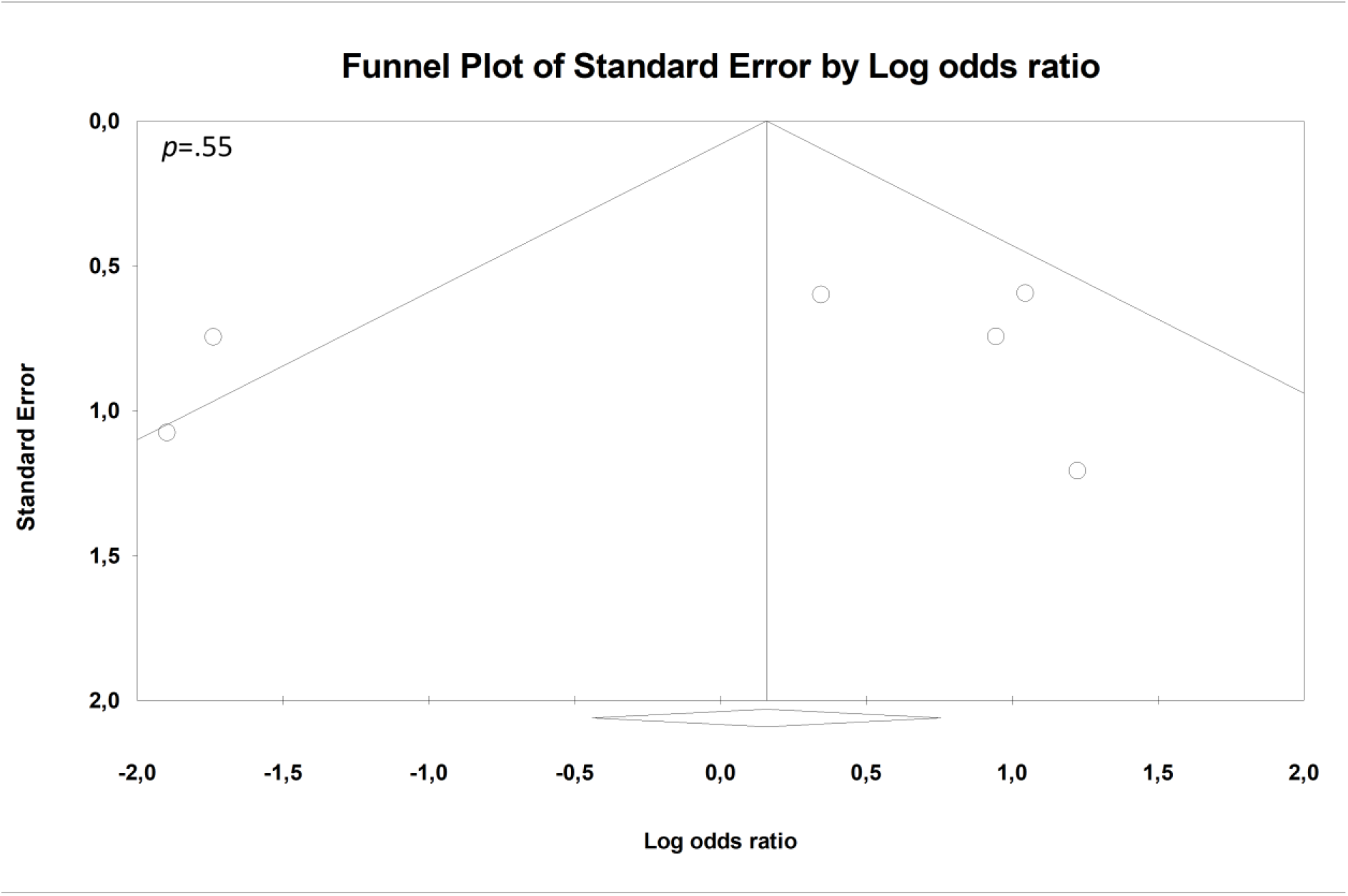
Funnel plot of the included studies for the outcome of complete wound closure. Studies were sparse and asymmetrical which indicates those studies had high risk of bias.

### 6.4 Inflammation/Infection

The presence of inflammation/infection was reported in 4 RCTs^6,7,10,22^ and 1 non-controlled interventional study^31^. Of the 4 RCTs, 2 studies (Wattanaploy *et al.*, 2017 & Beele *et al.*, 2010) were not included in the meta-analysis due to reporting zero events of inflammation/infection in both groups, therefore giving no weight in the analysis. The pooled analysis yields n=284 from Malizos *et al.* (2017)^6^ and Ceviker *et al.* (2015)^7^ studies.

The use of coating of osteosynthesis implants with antibiotic-loaded hydrogel (DAC®) in a study by Malizos *et al.* (2017) showed a significant difference between groups where the incidence of inflammation/infection was 0% (0 of 126) in the intervention group. In contrast, the control group exhibited an incidence of 4.7% (6 of 127).

The treatment of PHMB-based products as an irrigation solution and topical application in the study by Ceviker *et al.* (2015) demonstrated that after a 52-week observation, the infection rate on coronary bypass post-surgical wounds, as identified by culture, was 40% in the intervention group and 37.5% in the control group. However, there was no significant difference between the groups (p=0.886).

The pooled estimate of this outcome for those studies is LOD −0.95 (95% CI: −3.54 – 1.64), while the between-study heterogeneity variance was estimated at τ^2^=2.32 (Q=2.71, *p*=.10) (Figure 6).

**Figure 6.**
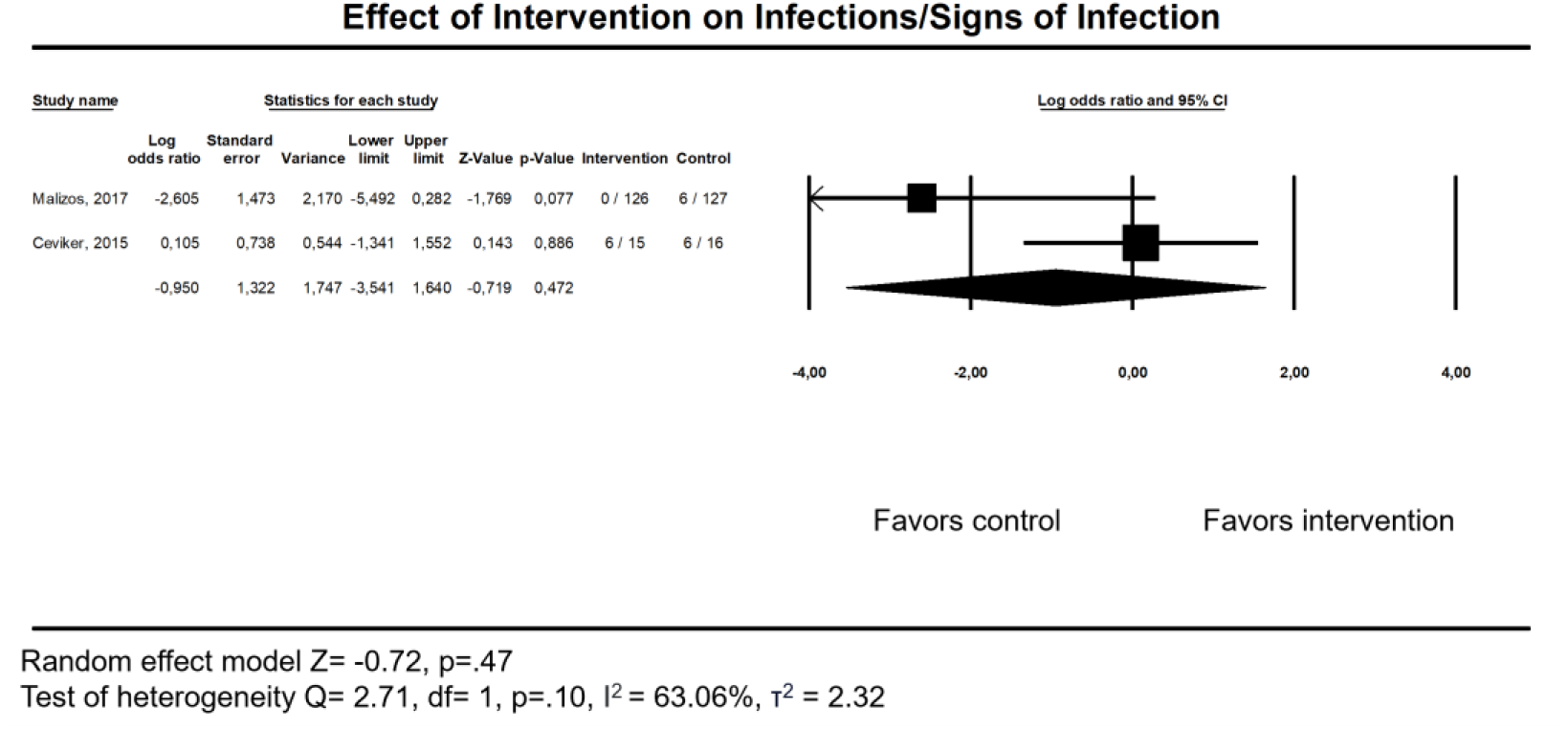
Forest plot of the effect of intervention on the presence of infection/signs of inflammation.

### 6.5 Bioburden

The effect of bacterial load reduction was reported in 4 RCTs^8,12,16,23^ and 5 case series or non-controlled interventional study^15,17,25,30,31^. However, the pooled analysis of the RCTs was not feasible due to poor data report (Borges *et al.,* 2018 & Kim *et al.,* 2018)^8,23^ and ununifiable outcomes (Yang *et al.,* 2017 & Malone *et al.,* 2019)^12,16^. The five case series were conducted by Gupta *et al.* (2019)^25^, Patel *et al.* (2021)^37^, Malone *et al.* (2021)^15^, Dryden *et al.* (2016)^30^, and Francesco *et al.* (2022)^31^.

Case series by Gupta *et al.* (2019)^25^ and Patel *et al.* (2021)^37^ studied the use of topical bacteriophage therapy isolated from different water sources (ponds, rivers, and sewers). Using a convenient sample of 20 and 48 chronic non-healing wounds, respectively, they showed that after a 2-week post-intervention, all wounds became sterile.^17,25^

Malone *et al.* (2021)^15^ studied the application of surfactant gel Plurogel®in addition to the standard of care and showed that 7 of 10 wounds had a Log10 reduction in bioburden.

Dryden *et al.* (2016)^30^ studied the application of Surgihoney RO™ (Matoke Holdings Ltd., UK), a pure honey-derived gel with enhanced reactive oxygen species, on acute and chronic ulcers. After four weeks of application, 98% of the wounds exhibited a bacterial load reduction as evaluated via semiquantitative culture, but no statistical significance estimation was provided.

While Francesco *et al.* (2022)^31^ utilized the topical application of hyaluronic acid in combination with silver sulfadiazine on complicated chronic wounds and showed a significant Log10 bacterial load reduction four weeks after the baseline.

## 7. Discussion

This systematic review and meta-analysis aimed to elucidate the effectiveness of various interventions in targeting biofilms and their impact on wound healing outcomes in clinical studies. According to our review, the effect of any intervention aimed at biofilm elimination and complete biofilm presence eradication could not be clarified due to the lack of clinical trials available. However, we could estimate its pooled effect on the secondary outcomes: complete wound closure and presence of inflammation/infection. Still, we found that all the products claimed to have anti-biofilm properties showed no significant effect in achieving complete wound closure and reducing inflammation/infection compared to the control treatment.

Clinical trials we found involving techniques of visual-guided wound cleansing, including wound-blotting guided debridement and autofluorescence imaging camera; and wound care products, including silver-based products, PHMB dressing and irrigation solution, NPWT with instillation of sodium hypochlorite, cadexomer iodine, and antibiotic-impregnated gels. One interesting approach is using topical bacteriophage cocktail studies by Gupta *et al.* (2019)^25^ and Patel *et al.* (2021)^37^in case series studies where the wound became sterile after the cocktail treatment. Although they did not evaluate the biofilm directly itself, this might suggest that the treatment might prevent the development of biofilm in the first place, showing the potential for further exploration in the future.

The problem with studies included in this review is that there needed to be more consensus on objectively evaluating the biofilm. Studies included in this review still rely on the conventional culture method or surrogate end-point, such as clinical signs and symptoms. Studies by Beele *et al.* (2010) use clinical signs and symptoms to identify biofilms, such as continuous pain, edema, warmth, moderate to heavy exudate, and the presence of slough at least 50% of the wound surface, foul odor, or necrotic plaques. Studies by Yang *et al.* (2017)^16^, Miller *et al.* (2010)^9^, and Wolcott *et al.* (2015)^24^, were also claiming the evaluation of intervention on the biofilm matrix. However, these studies needed to present data regarding the objective evaluation of the biofilm’s presence. Instead, they focused on the colony-forming unit count and explored its influence on wound healing. Only studies by Borges *et al.* (2018)^8^ and Malone *et al.* (2019 & 2021)^12,15^ reported objective evaluation of biofilm using scanning electron microscopy, DNA sequencing, and real-time quantitative polymerase chain reaction (qPCR). Yet this technique was used only to confirm the biofilm presence at the baseline, making the quantitative evaluation at the end of the intervention not possible. This finding alarms the need of consensus for the objective biofilm evaluation for future studies.

Using a visual-guided wound cleansing approach may offer a solution in the clinician’s best interest more objectively. Techniques such as wound-blotting and autofluorescence-guided imaging may offer a promising approach for assessing biofilm presence and monitoring the effectiveness of interventions in real-time. The wound-blotting technique could visualize the wound surface biofilm by attaching a piece of nitrocellulose membrane on wounds for 10 seconds, then stained by Alcian Blue to highlight parts of the wound that still harbor biofilms. This technique has a sensitivity of 100%, as shown in *in vivo* study.^38^ An RCT by Astrada *et al.* (2021) and observational studies by Mori *et al.* (2019)^21^ and Koyanagi *et al*. (2021)^20^ show that wound blotting could help clinicians to determine the effectiveness of wound cleansing in real-time as well as improving wound healing. While in The Fluorescence imaging Assessment and Guidance (FLAAG) study, a multi-center diagnostic accuracy study by Le *et* al. (2022)^39^ on 350 patients with chronic wounds, shows that the autofluorescence imaging technique could detect >10^4^ CFU/g in 82% of biopsied wound tissue which mostly missed by direct inspection of the clinical signs and symptoms. The study shows this technique has moderate sensitivity (61,0%) but high specificity (84%).^39^ A study by Rahma *et al.* (2022)^14^ is the first RCT to evaluate this technique, but it is still a pilot study. Regardless, this technique could change 69% of treatment plans, influenced 85% of wound bed preparation, and improved 90% of patient care by 20 clinicians in those centers.^39^ Additionally, a study by Wu *et al.* (2022)^32^ attempted to compare both techniques, i.e., wound-blotting and autofluorescence imaging, in predicting healing in chronic wounds in 90 days. They found that wound blotting shows a significantly strong correlation coefficient of Kendall’s tau value = 0.563, p<.001 to complete wound healing, while MolecuLight i:X Exhibited no significant association (p=.184). This indicates that wound blotting may be more beneficial for wound healing prognosis because the wound blotting detects the actual biofilms. In contrast, the autofluorescence bacterial visualization detects planktonic and biofilm forms but cannot distinguish between the two.^38^ Future research is needed to focus on standardized methods for biofilm assessment and to assess the long-term effects of biofilm elimination on wound healing outcomes considering the clinical implications of biofilm elimination in improving patient well-being.

Finally, since we found no sufficient evidence supporting the antibiofilm effects of any available products or topical interventions, the skills of wound care clinicians seem to be the most critical factor in biofilm management. This may include the cleansing technique, locating the highly susceptible wound bed for biofilm development, exudate management, antimicrobial selection, and adherence to the timely dressing change. Visual-guided wound cleansing may open a new therapeutic approach to combat wound surface biofilm.

### Limitations

There is a high risk of bias in almost all clinical trials included in the meta-analysis. The primary source of bias is the lack of consensus on objectively evaluating the biofilm. There is also the risk of selection bias because studies included in the pooled estimation did not distinguish types of modalities due to the lack of eligible clinical trials.

## 8. Conclusion

This study indicates that there is currently inadequate evidence to support the claims of any topical treatments having an anti-biofilm effect on the healing of acute and chronic wounds. The skill of treating clinicians offers the main contribution in eliminating biofilm and improving wound healing which can be optimized via visual-guided wound cleansing such as wound blotting and autofluorescence imaging techniques. More rigorous clinical trial studies are needed to clarify the benefit of those techniques.

## 9. Take-Home Messages

- Insufficient Clinical Backing for Anti-Biofilm Treatments: Despite claims, this analysis highlights the scarcity of clinical trials supporting biofilm-targeting interventions in wound healing, necessitating more substantial evidence.
- Key Clinician Skill in Biofilm Management: The proficiency of wound care practitioners stands pivotal in biofilm control and wound healing, encompassing techniques, antimicrobial choices, and dressing adherence.
- Visual-Guided Cleansing Shows Promise: Techniques like wound-blotting and autofluorescence imaging offer potential in objectively assessing biofilm presence, influencing real-time treatment decisions, and warranting further study for enhanced wound healing.

## Data Availability

All data produced in the present work are contained in the manuscript

## 10. Authors’ Contributions

AA contributed to the design of this review. AA and KRA collect all data and screen data. AA and RAP contributed to data verification and statistical analysis. AA made all figures and tables. AA, RAP, and KRA contributed to the article writing. AA and RAP revised the article. All listed authors checked and agreed to the final article.

## 11.#Acknowledgement and Funding

This research received no specific grant from funding agencies in the public, commercial, or not-for-profit sectors.

## 12. Author Disclosure and Ghostwriting

The authors declare that no conflict of interest might affect the integrity of this work. While preparing this work, the authors used ChatGPT August 3 Version to improve the readability of the text. After using this tool/service, the authors reviewed and edited the content as needed and take full responsibility for the publication’s content.

## 13. About the Authors

**Adam Astrada**, an assistant professor at Esa Unggul University and a national board member of the Indonesian Wound Enterostomal and Continence Nurses Association, who has received a Gold Prize from the Journal of Wound Care Awards 2023 for his works in Advances in Infections and Biofilm. He has published 4 SCI, including 3 articles in the Journal of Wound Care and 1 article in Enfermeria Clinica.

**Rian Adi Pamungkas**, an assistant professor and Head of the School of Nursing at Esa Unggul University. He is also a postdoctoral scholar at the University of Michigan.

**Khoirul Rista Abidin** is a doctoral student in Biotechnology Study Program, at Padjadjaran University. He is a Lecturer at the Department of Medical Laboratory Technology, Aisyiyah Polytechnic Pontianak. He also works as a wound care clinician at Komamura Wound & Obesity Care Center and Aisyiyah Clinic Pontianak. His research interest is noninfectious diseases.

## 14. Abbreviations and Acronyms

CI: confidence interval
DAC: defensive antibacterial coating
DFUs: diabetic foot ulcers
DMIST: depth, maceration, inflection/inflammation, size, tissue of wound bed, tissue of wound edge, and tunneling/undermining
LOD: log odds ratio
MeSH: medical subject headings
NaCl: natrium chloride
NPWT: negative pressure wound therapy
PFU: plaque forming unit
PHMB: poly-hexamethylene biguanide
PRISMA: preferred reporting items for systematic reviews and meta-analyses
RCTs: randomized controlled trials
SD: standard deviation SE: standard error
SOC: standard of care
TBSA: total body surface area

## 16. Appendix 1

### 16.1 Search Strategies

#### PubMed - run 17/8/2022

(““dressing*““[Text Word] OR ““occlusive dressing*““[MeSH Terms] OR ““biological dressing*““[MeSH Terms] OR ““biological dressing*““[Text Word] OR ““bandage*““[Text Word] OR ““bandage*““[MeSH Terms] OR (““ointment*““[Text Word] OR ““topical ointment*““[Text Word] OR ““anti infective agent*““[MeSH Terms] OR ““topical*““[Text Word] OR ““mupirocin*““[MeSH Terms] OR ““oxyquinolin*““[MeSH Terms] OR ““sulfacetamide*““[MeSH Terms] OR ““silver sulfadiazine*““[MeSH Terms] OR ““silver protein*““[MeSH Terms] OR ““proflavine*““[MeSH Terms] OR ““povidone iodine*““[MeSH Terms] OR ““natamycin*““[MeSH Terms] OR ““ointment*““[MeSH Terms] OR ““nitrofurazone*““[MeSH Terms] OR ““miconazole*““[MeSH Terms] OR ““hydroxyquinoline*““[MeSH Terms] OR ““hydrogen peroxide*““[MeSH Terms] OR ““hexylresorcinol*““[MeSH Terms] OR ““ethacridin*““[MeSH Terms] OR ““econazole*““[MeSH Terms] OR ““chlorhexidine*““[MeSH Terms] OR ““cetylpyridin*““[MeSH Terms] OR ““camphor*““[MeSH Terms] OR ““benzethonium*““[MeSH Terms])) AND (““antibiofilm*““[Text Word] OR ““biofilm*““[Text Word] OR ““biofilm*““[MeSH Terms] OR ““extracellular polymeric substance matri*““[MeSH Terms]) AND (““wound*““[Text Word] OR ““wounds and injuries““[MeSH Terms] OR ““wound infection““[MeSH Terms] OR ““wound healing““[MeSH Terms] OR ““surgical wound infection““[MeSH Terms] OR ““surgical wound““[MeSH Terms] OR ““ulcer*““[Text Word] OR ““foot ulcer*““[MeSH Terms] OR ““skin ulcer*““[MeSH Terms] OR ““leg ulcer*““[MeSH Terms] OR ““injur*““[Text Word] OR ““skin injur*““[Text Word] OR ““skin injur*““[Text Word])

## Notes

### Competing Interest Statement

The authors have declared no competing interest.

### Funding Statement

This study did not receive any funding

## References

1. National Health Institute. Research on Microbial Biofilms. 2002. Available from: https://grants.nih.gov/grants/guide/pa-files/pa-03-047.html [Last accessed: 9/3/2022].

2. Yan J, Bassler BL. Surviving as a Community: Antibiotic Tolerance and Persistence in Bacterial Biofilms. Cell Host Microbe 2019;26(1):15–21; doi: 10.1016/j.chom.2019.06.002.

3. Hrynyshyn A, Simões M, Borges A. Biofilms in Surgical Site Infections: Recent Advances and Novel Prevention and Eradication Strategies. Antibiotics 2022;11(1):69; doi: 10.3390/antibiotics11010069.

4. Malone M, Bjarnsholt T, Mcbain AJ, et al. The prevalence of biofilms in chronic wounds: a systematic review and meta-analysis of published data. J Wound Care 2017;26(1):20–25.

5. Schwarzer S, James GA, Goeres D, et al. The efficacy of topical agents used in wounds for managing chronic biofilm infections: A systematic review. J Infect 2020;80(3):261–270; doi: 10.1016/j.jinf.2019.12.017.

6. Malizos K, Blauth M, Danita A, et al. Fast-resorbable antibiotic-loaded hydrogel coating to reduce post-surgical infection after internal osteosynthesis: a multicenter randomized controlled trial. J Orthop Traumatol 2017;18(2):159– 169; doi: 10.1007/s10195-017-0442-2.

7. Ceviker K, Canikoglu M, Tatlioglu S, et al. Reducing the pathogen burden and promoting healing with polyhexanide in non-healing wounds: A prospective study. J Wound Care 2015;24(12):582–586; doi: 10.12968/jowc.2015.24.12.582.

8. Borges EL, Frison SS, Honorato-Sampaio K, et al. Effect of Polyhexamethylene Biguanide Solution on Bacterial Load and Biofilm in Venous Leg Ulcers. J Wound, Ostomy Cont Nurs 2018;45(5):425–431; doi: 10.1097/WON.0000000000000455.

9. Miller CN, Newall N, Kapp SE, et al. A randomized-controlled trial comparing cadexomer iodine and nanocrystalline silver on the healing of leg ulcers. Wound Repair Regen 2010;18(4):359–367; doi: 10.1111/j.1524-475X.2010.00603.x.

10. Beele H, Meuleneire F, Nahuys M, et al. A prospective randomised open label study to evaluate the potential of a new silver alginate/carboxymethylcellulose antimicrobial wound dressing to promote wound healing. Int Wound J 2010;7(4):262–270; doi: 10.1111/j.1742-481X.2010.00669.x.

11. Harding KG, Szczepkowski M, Mikosiński J, et al. Safety and performance evaluation of a next-generation antimicrobial dressing in patients with chronic venous leg ulcers. Int Wound J 2016;13(4):442–448; doi: 10.1111/iwj.12450.

12. Malone M, Schwarzer S, Radzieta M, et al. Effect on total microbial load and community composition with two vs six-week topical Cadexomer Iodine for treating chronic biofilm infections in diabetic foot ulcers. Int Wound J 2019;16(6):1477–1486; doi: 10.1111/iwj.13219.

13. Astrada A, Nakagami G, Kashiwabara K, et al. Targeting Biofilms in the Management of Diabetic Foot Ulcers through Wound Blotting-Guided Visualization (ウンドブロッティングによる可視化に基づく バイオフィルムをターゲットとした糖尿病足潰瘍管理). The University of Tokyo; 2021.

14. Rahma S, Woods J, Brown S, et al. The Use of Point-of-Care Bacterial Autofluorescence Imaging in the Management of Diabetic Foot Ulcers: A Pilot Randomized Controlled Trial. Diabetes Care 2022;45(7):1601–1609; doi: 10.2337/dc21-2218.

15. Malone M, Radzieta M, Schwarzer S, et al. Efficacy of a topical concentrated surfactant gel on microbial communities in non-healing diabetic foot ulcers with chronic biofilm infections: A proof-of-concept study. Int Wound J 2021;18(4):457–466; doi: 10.1111/iwj.13546.

16. Yang C, Goss SG, Alcantara S, et al. Effect of Negative Pressure Wound Therapy With Instillation on Bioburden in Chronically Infected Wounds. Wounds a Compend Clin Res Pract 2017;29(8):240–246.

17. Patel DR, Bhartiya SK, Kumar R, et al. Use of Customized Bacteriophages in the Treatment of Chronic Nonhealing Wounds: A Prospective Study. Int J Low Extrem Wounds 2021;20(1):37–46; doi: 10.1177/1534734619881076.

18. Albaugh KW, Biely SA, Cavorsi JP. The effect of a cellulose dressing and topical vancomycin on methicillin-resistant Staphylococcus aureus (MRSA) and Gram-positive organisms in chronic wounds: a case series. Ostomy Wound Manage 2013;59(5):34–43.

19. Liu J, Cai M, Yan H, et al. Yunnan baiyao reduces hospital-acquired pressure ulcers via suppressing virulence gene expression and biofilm formation of staphylococcus aureus. Int J Med Sci 2019;16(8):1078–1088; doi: 10.7150/ijms.33723.

20. Koyanagi H, Kitamura A, Nakagami G, et al. Local wound management factors related to biofilm reduction in the pressure ulcer: A prospective observational study. Japan J Nurs Sci 2021;18(2):1–8; doi: 10.1111/jjns.12394.

21. Mori Y, Nakagami G, Kitamura A, et al. Effectiveness of biofilm-based wound care system on wound healing in chronic wounds. Wound Repair Regen 2019;27(5):540–547; doi: 10.1111/wrr.12738.

22. Wattanaploy S, Chinaroonchai K, Namviriyachote N, et al. Randomized controlled trial of polyhexanide/betaine gel versus silver sulfadiazine for partial-thickness burn treatment. Int J Low Extrem Wounds 2017;16(1):45–50; doi: 10.1177/1534734617690949.

23. Kim D, Namen Ii W, Moore J, et al. Clinical Assessment of a Biofilm-disrupting Agent for the Management of Chronic Wounds Compared With Standard of Care: A Therapeutic Approach. Wounds a Compend Clin Res Pract 2018;30(5):120–130.

24. Wolcott R. Disrupting the biofilm matrix improves wound healing outcomes. J Wound Care 2015;24(8):366–371; doi: 10.12968/jowc.2015.24.8.366.

25. Gupta P, Singh HS, Shukla VK, et al. Bacteriophage Therapy of Chronic Nonhealing Wound: Clinical Study. Int J Low Extrem Wounds 2019;18(2):171– 175; doi: 10.1177/1534734619835115.

26. Walker M, Metcalf D, Parsons D, et al. A real-life clinical evaluation of a next-generation antimicrobial dressing on acute and chronic wounds. J Wound Care 2015;24(1); doi: 10.12968/jowc.2015.24.1.11.

27. Lenselink E, Andriessen A. A cohort study on the efficacy of a polyhexanide-containing biocellulose dressing in the treatment of biofilms in wounds. J Wound Care 2011;20(11):534–539; doi: 10.12968/jowc.2011.20.11.534.

28. Metcalf DG, Bowler PG. Clinical impact of an anti-biofilm hydrofiber dressing in hard-to-heal wounds previously managed with traditional antimicrobial products and systemic antibiotics. Burn Trauma 2020;8:1–9; doi: 10.1093/BURNST/TKAA004.

29. Metcalf D, Parsons D, Bowler P. A next-generation antimicrobial wound dressing: a real-life clinical evaluation in the UK and Ireland. J Wound Care 2016;25(3):132–138; doi: 10.12968/jowc.2016.25.3.132.

30. Dryden M, Dickinson A, Brooks J, et al. A multi-centre clinical evaluation of reactive oxygen topical wound gel in 114 wounds. J Wound Care 2016;25(3):140–146; doi: 10.12968/jowc.2016.25.3.140.

31. De Francesco F, Riccio M, Jimi S. Contribution of Topical Agents such as Hyaluronic Acid and Silver Sulfadiazine to Wound Healing and Management of Bacterial Biofilm. Medicina (B Aires) 2022;58(6):835; doi: 10.3390/medicina58060835.

32. Wu Y, Lin Y, Yang H, et al. Point-of-Care Wound Blotting with Alcian Blue Grading versus Fluorescence Imaging for Biofilm Detection and Predicting 90-Day Healing Outcomes. Biomedicines 2022;10(5):1200; doi: 10.3390/biomedicines10051200.

33. Romanelli M, Dini V, Barbanera S, et al. Evaluation of the efficacy and tolerability of a solution containing propyl betaine and polihexanide for wound irrigation. Skin Pharmacol Physiol 2010;23(SUPPL. 1):41–44; doi: 10.1159/000318266.

34. Metcalf D, Parsons D, Bowler P. A next-generation antimicrobial wound dressing: a real-life clinical evaluation in the UK and Ireland. J Wound Care 2016;25(3):132–138; doi: 10.12968/jowc.2016.25.3.132.

35. Mori Y, Nakagami G, Kitamura A, et al. Effectiveness of biofilm-based wound care system on wound healing in chronic wounds. Wound Repair Regen 2019;27(5):540–547; doi: 10.1111/wrr.12738.

36. Oe M, Yotsu RR, Arisandi D, et al. Validity of DMIST for monitoring healing of diabetic foot ulcers. Wound Repair Regen 2020;28(4):539–546; doi: 10.1111/wrr.12816.

37. Serena TE, Jalodi O, Serena L, et al. Evaluation of the combination of a biofilm-disrupting agent and negative pressure wound therapy: A case series. J Wound Care 2021;30(1):9–14; doi: 10.12968/jowc.2021.30.1.9.

38. Astrada A, Nakagami G, Minematsu T, et al. Concurrent validity of biofilm detection by wound blotting on hard-to-heal wounds. J Wound Care 2021;30(Sup4):S4–S13; doi: 10.12968/jowc.2021.30.Sup4.S4.

39. Le L, Baer M, Briggs P, et al. Diagnostic Accuracy of Point-of-Care Fluorescence Imaging for the Detection of Bacterial Burden in Wounds: Results from the 350-Patient Fluorescence Imaging Assessment and Guidance Trial. Adv Wound Care 2021;10(3):123–136; doi: 10.1089/wound.2020.1272.

40. Miller CN, Carville K, Newall N, et al. Assessing bacterial burden in wounds: comparing clinical observation and wound swabs. Int Wound J 2011;8(1):45–55; doi: 10.1111/j.1742-481X.2010.00747.x.

41. Metcalf DG, Bowler PG. Clinical impact of an anti-biofilm Hydrofiber dressing in hard-to-heal wounds previously managed with traditional antimicrobial products and systemic antibiotics. Burn Trauma 2020;8:1–9; doi: 10.1093/burnst/tkaa004.

